# HCLmNet: A Unified Hybrid Continual Learning Strategy Multimodal Network for Lung Cancer Survival Prediction

**DOI:** 10.1101/2024.12.14.24319041

**Authors:** Ilias Bappi, David J. Richter, Shivani Sanjay Kolekar, Kyungbaek Kim

## Abstract

Lung cancer survival prediction is a critical task in healthcare, where accurate and timely predictions can significantly impact patient outcomes. In hospital settings, new patient data is constantly generated, requiring predictive models to adapt without forgetting previously learned knowledge. This challenge is intensified by the need to seamlessly integrate complex multimodal data, such as imaging, DNA, and patient records. Traditional Deep Learning (DL) models, while powerful, often suffer from catastrophic forgetting during incremental learning, further complicating the task of reliable survival prediction in dynamic environments. To address these challenges, we introduce a hybrid Continual Learning (CL) framework that integrates Elastic Weight Consolidation (EWC) with replay-based modules, including EWC Experience Replay (ER), Instance-Level Correlation Replay (EICR), and Class-Level Correlation Replay (ECCR). The ER module preserves knowledge by replaying representative samples from previous data, mitigating interference from new data. The EICR module ensures the retention of fine-grained feature patterns through inter-instance relationship modeling, while the ECCR module consolidates global knowledge across tasks using random triplet probabilities to preserve inter-class correlations. Together, these components create a robust framework, addressing catastrophic forgetting while enhancing adaptability for real-time survival prediction. Another critical challenge is the limitations of Convolutional Neural Networks (CNNs), which tend to miss ground-glass opacities or tiny tumor features in CT and PET images due to their reliance on datasets similar to their pretraining data. To overcome this, we propose a Swin Transformer (SwinT)-based method to extract critical features, addressing CNN shortcomings in such multimodal scenarios. Additionally, XLNet-permutation enriches multimodal analysis by effectively handling small DNA datasets and capturing latent patterns, whereas Fully Connected Network (FCN) process clinical features. A cross-attention fusion mechanism integrates clinical, CT, PET, and DNA data, producing a robust survival prediction model. The final prediction is guided by FCN and Cox Proportional Hazards (CoxPH) techniques, achieves state-of-the-art performance with a 7.7% concordance index (C-Index) improvement (0.84), a mean absolute error (MAE) reduction to 140 days, and minimized forgetting to 0.08. Ablation studies demonstrate the importance of the DNA modality, cross-attention mechanism, and CL strategies, advancing adaptive survival prediction and stability.

## 1 Introduction

Lung cancer is one of the most common malignancies worldwide and accounts for 18% of cancer-related deaths [1]. In the field of lung cancer survival analysis medical data such as clinical (patient observation data), Computed tomography (CT), Positron emission tomography (PET), and DNA genomic mutation single nucleotide variants(SNV), Heterozygous (HETE), and Homozygous (HOMO) is essential for the early detection, monitoring, diagnosis and treatment of this disease.Traditionally, radiologists and medical experts rely on visual inspection of medical images to identify tumor instances or other relevant factors, often supplemented by patient history and experimental records. This process is time-intensive and prone to inaccuracies, as survival time predictions and treatment decisions are frequently inftiluenced by clinicians subjective knowledge and experience. While DL models have been employed to improve prediction accuracy, they face notable challenges. CNNs, for instance, struggle with detecting small or multiple tumor instances in high-resolution medical images [2, 3]. Additionally, the integration of heterogeneous data sources, such as clinical, imaging, and genomic datasets, further complicates survival prediction due to differences in feature representation.

An equally critical challenge in survival prediction lies in the application of CL, where models must incorporate new data without overwriting prior knowledge a phenomenon termed catastrophic forgetting [4]. CL methods have emerged as a promising solution, enabling models to maintain and adapt existing knowledge while processing new tasks [5]. Existing CL strategies can be broadly categorized into regularization-based methods [6, 7], structure-based approaches [8–11], and replay-based techniques [12–14]. Replay-based methods, in particular, mimic human learning by selectively storing and reviewing past experiences, offering a computationally efficient way to alleviate forgetting. Classical replay techniques [13], such as Experience Replay, have demonstrated impressive performance by replaying a mix of previous and current tasks to reinforce learned patterns. However, conventional methods often overlook critical aspects, such as preserving the instance-level and class-level correlations essential for maintaining structural consistency across tasks. Additionally, when the new data are large or complex, replay-based methods with memory buffers alone can fall short. In such cases, EWC offers a complementary solution, particularly when integrated with replay-phase strategies.

To address these gaps, this study proposes a hybrid CL framework combining EWC with replay-based modules: ER, EICR, and ECCR. Each module serves a distinct purpose: ER ensures the retention of fundamental knowledge by replaying a mix of past and current tasks, promoting gradual and stable learning, EICR maintains consistency at the instance level by constructing a correlation matrix that captures inter-instance relationships, preserving structural information critical to individual data points, and ECCR reinforces class-level consistency by leveraging contrastive learning principles, particularly using random triplet mechanisms (anchor, positive, and negative) to maintain clear boundaries between classes, ensuring distinctions like tumor and non-tumor regions are preserved over time. This combination enables a robust mechanism to mitigate catastrophic forgetting while ensuring scalability for complex data.

Our approach integrates these strategies with FCN for maintaining clinical features sequecnces with other modalaties and SwinT for feature extraction, enhancing the detection of critical tumor features such as ground-glass opacities and small tumor instances from CT and PET images. Moreover, XLNet-permutation is employed to effectively handle small DNA datasets, uncovering latent genomic patterns that enrich multimodal survival prediction. Finally, a cross-attention fusion mechanism integrates clinical, CT, PET, and DNA data, ensuring comprehensive and robust survival predictions via CoxPH modeling. The overall framework is shown in Fig 2. In summary, there are primary highlights of our research:

- We introduced a hybrid CL framework that integrates EWC with Replay-phase mechanisms, addressing catastrophic forgetting and enabling dynamic adaptation for lung cancer survival prediction.
- We developed three complementary replay and EWC-based mechanisms ER, EICR, and ECCR within the hybrid framework. ER retains fundamental knowledge, EICR preserves inter-instance relationships, and ECCR maintains robust class-level boundaries, collectively enhancing scalability for large or complex datasets.
- We employed a SwinT-based feature extraction method that significantly improves the detection of critical lung cancer features, including ground-glass opacities and small tumor instances in CT and PET scans.
- Leveraged XLNet-permutation to effectively process small DNA datasets, uncovering latent genomic patterns and enriching the multimodal survival prediction framework.
- Inspired by cross-attention fusion techniques, we developed a novel cross-attention strategy to integrate clinical, CT, PET, and DNA data, ensuring comprehensive survival prediction using CoxPH modeling with real-time adaptability through CL techniques.
- We conducted a thorough ablation study and comparative evaluation with state-of-the-art models, demonstrating the superior performance and robustness of the proposed hybrid CL-based multimodal framework.

This paper is organized as follows: Section 2 reviews related work in survival prediction and continual learning. Section 3 discusses this research background. Section 4 describes the proposed framework and methodological innovations. Experimental setup are presented in Section 5, Experimental results and evaluations are presented in Section 6, followed by conclusions and future directions in Section 7.

## 2 Related work

### Lung Cancer in survival prediction

Survival prediction for lung cancer is critical for guiding personalized treatment strategies and improving patient outcomes. Lung cancer research focuses on predicting survival time, recommending optimal treatments, and providing comprehensive prognosis insights based on clinical and imaging data. A notable advancement in this domain was made by Sesen et al. [15], who utilized the LUCADA dataset to develop a framework for predicting 1-year survival rates while simultaneously recommending treatment plans for lung cancer patients. Building on this, Yu et al. [16] improved survival prediction by modeling survival distributions as a series of dependent tasks. They used sequential regressors to capture the temporal relationships in survival data, thus enhancing the accuracy of predictions. Paul et al. [17] further demonstrated the potential of deep learning in survival prediction by extracting features from CT images of lung cancer patients using CNNs and applying a nearest neighbor classifier for survival estimation. This integration of imaging features has paved the way for leveraging high-resolution imaging data in survival prediction tasks. Similarly, advancements in combining imaging with multimodal datasets have strengthened survival prediction frameworks, as demonstrated in broader studies addressing cancer survivability [18]. While these works emphasize the importance of data-driven modeling in lung cancer survival prediction, they also reflect the challenges of integrating diverse data types and maintaining consistency in predictions across heterogeneous datasets. The continued refinement of imaging techniques, multimodal integration, and personalized modeling is essential for improving the clinical applicability of survival prediction frameworks in lung cancer.

### Advancements in Deep Learning for Survival Prediction

DL has emerged as a transformative approach in survival prediction, providing enhanced capabilities for handling complex medical data. Traditional survival models like Cox regression often struggle to capture nonlinear relationships in high-dimensional data. To address this limitation, neural networks have been adapted to optimize Cox’s negative partial likelihood, leading to improved or comparable performance in survival predictions, particularly when analyzing challenging medical imaging data [19, 20]. Recent advancements include CNN-based survival models that integrate DL with clinical outcomes. For example, Zhu et al. introduced DeepConvSurv, a model that combines CNNs with Cox regression to predict survival from regions of interest (ROIs) in lung cancer histology images, offering hazard rate predictions at a patch level [21]. Similarly, Mobadersany et al. developed Survival CNNs, leveraging high-power fields (HPFs) from ROIs to provide patient-level survival predictions by aggregating median risks [22]. Further enhancing these techniques, Yao et al. proposed Deep Attention Multiple Instance Survival Learning (DeepAttnMISL), utilizing attention-based aggregation for whole-slide image (WSI) feature learning [23]. This approach addressed the limitations of patch-level predictions by clustering WSI patches into more meaningful groupings, ensuring robust patient-level hazard rate estimation.

Building on these DL advancements, techniques such as Multiple Instance Learning (MIL) have further enhanced survival prediction, especially in weakly supervised settings where only patient-level labels are available. MIL frameworks often employ CNNs to generate instance-level embeddings, followed by aggregation networks that compute bag-level predictions. For instance, Wang et al. applied recalibrated MIL to gastric cancer classification [24], while Liu et al. utilized landmark-based MIL for brain disease diagnosis [25]. The integration of MIL with DL has enhanced survival modeling by facilitating the analysis of large-scale datasets and enabling the aggregation of complex patterns from ROIs or patches. Aggregation methods in MIL have evolved from naive approaches, such as averaging or selecting extreme hazard rates, to more sophisticated trainable techniques. Non-trainable methods, while computationally simple, fail to capture intricate interrelations between patches. In contrast, trainable aggregation methods, such as RNN-based and attention-based techniques [24, 26, 27], have shown superior performance in representing the survival function by learning the underlying relationships within the data. Despite significant advancements, existing DL-based frameworks often struggle with the effective integration of multimodal data, such as clinical, imaging, and genomic information. To address these challenges, our proposed multimodal model leverages FCN for clinical data, SwinT for imaging data, XLNet for genomic data, and a cross-attention mechanism to facilitate seamless integration across modalities, followed by FCN refinement within the CoxPH framework for prediction. While these innovations enhance survival predictions, existing models face persistent difficulties in adapting to continuously evolving datasets without overwriting prior knowledge. This limitation highlights the critical need for CL strategies, which are discussed in the next section.

### Continual Learning in Survival Prediction

CL focuses on the progressive acquisition of knowledge from an ever-growing stream of data while retaining previously learned information. This capability is vital in real-world scenarios, where survival prediction models must adapt to evolving datasets without compromising the knowledge gained from prior tasks [4, 28, 29]. However, a significant challenge in CL is catastrophic forgetting, where neural networks tend to lose previously acquired knowledge when trained on new tasks. To address this issue, researchers have developed several CL strategies, which can be broadly categorized into regularization-based, structure-based, and replay-based methods.

(i)Regularization-Based Methods: These approaches mitigate forgetting by constraining changes to model parameters during new task learning. Methods like EWC [30] introduce a quadratic penalty to balance parameter updates for old and new tasks. Similarly, Synaptic Intelligence [31] identifies parameters critical to previous tasks and penalizes their changes. Learning without Forgetting [7] employs knowledge distillation to transfer knowledge into a smaller model while retaining prior information. (ii) Structure-Based Methods: These methods allocate specific model components to individual tasks, isolating knowledge to prevent interference. Progressive Neural Networks [8], for example, introduce separate networks for new tasks while leveraging earlier models for assistance. Although effective in reducing forgetting, these methods demand significant computational and memory resources due to the need for additional storage of task-specific parameters [9, 11, 32]. (iii) Replay-Based Methods: Replay-based methods alleviate forgetting by selectively replaying stored samples from previous tasks during training. Approaches like Experience Replay [13] combine stored and current data in mini-batches to train the model. Extensions like Dark Experience Replay [14] incorporate knowledge distillation to preserve the consistency of model logits over time. Variants such as Gradient Sample Selection [33] and Hard Anchor Learning [34] enhance the rehearsal process by prioritizing critical samples or introducing anchoring objectives to retain key information. Unlike conventional methods that rely solely on replay strategies, our approach overcomes the limitations posed by large-scale data. As memory size grows, traditional replay methods become less efficient and scalable. By incorporating EWC, we mitigate this issue through selective parameter regularization, reducing dependency on memory buffer size while enhancing knowledge retention.

To address the unique challenges of survival prediction in multimodal data, we propose a hybrid CL framework that combines the strengths of EWC and replay strategies. This framework introduces three key modules ER, EICR, and ECCR—to enhance adaptability and robustness. The ER Module merges EWC’s parameter-regularization capabilities with replay-based sample rehearsals. It calculates the total loss as the sum of the standard loss for new data and the EWC loss, which penalizes changes to parameters deemed important by the Fisher Information Matrix (FIM). This mechanism balances the retention of previous knowledge and the integration of new information. The EICR Module emphasizes maintaining instance-level consistency by constructing a correlation matrix that captures inter-instance relationships within the replay buffer. This matrix encodes structural information critical to preserving the alignment of feature representations of individual samples across tasks. The loss function is designed to penalize deviations from these relationships during incremental learning, ensuring fine-grained control over forgetting at the instance level. Additionally, the ECCR Module preserves class-level interactions by leveraging triplet-based learning. For each sample, an anchor, a positive (from the same class), and a negative (from a different class) are selected. The triplet loss ensures that the anchor-positive pair is pulled closer together while the anchor-negative pair is pushed further apart in the feature space. This mechanism not only maintains clear class boundaries, such as distinguishing tumor from non-tumor regions, but also reinforces the model’s understanding of inter-class relationships over time.

Each module (ER, EICR, and ECCR) integrates seamlessly into the overarching process, which operates through four critical phases: training, replay, prediction, and memory buffering. These phases collectively ensure the model’s ability to adapt to new datasets while preserving prior knowledge. By addressing the limitations of existing CL frameworks with advanced loss mechanisms tailored to each module, our hybrid approach fosters robust and adaptive survival prediction models designed for the complexities of multimodal data.

## 3 Background

Since lung cancer is one of the leading causes of cancer related deaths worldwide, early and precise survival projections are crucial for enhancing patient outcomes and clinical judgment. These forecasts are essential for directing treatment decisions and improving care plans. A crucial component of this process is the TNM staging system, developed by the American Joint Committee on Cancer (AJCC) and the International Union Against Cancer (UICC) [35]. This globally recognized system assesses the severity and spread of cancer in the body, where T describes the size of the tumor, N represents the spread to nearby lymph nodes, and M indicates metastasis to other body parts. In addition to TNM staging, other patient-specific attributes such as gender, age, smoking status and amount, survival time, and overall clinical stage play a critical role in survival prediction. Together, these multimodal data points offer a comprehensive view of a patient’s condition, enabling clinicians to make informed decisions about treatment strategies. However, leveraging these diverse data types to predict patient survival poses significant challenges in dynamic hospital environments. New patient data, including imaging, DNA, and updated medical records, is generated daily, requiring predictive models to adapt continually without losing prior knowledge. Conventional models for DL, while effective in processing complex datasets, often struggle with catastrophic forgetting where learning from new data overwrites previously acquired knowledge. This limitation undermines the reliability of survival predictions and hampers the integration of multimodal data in rapidly evolving clinical settings. To address this, hybrid CL strategies have emerged as a promising solution. Replay-based methods, which use memory buffers to retain critical information from previous data, are particularly effective for incremental learning. However, replay-based approaches alone may not be sufficient when the new data is big or complicated, such as when imaging, DNA profiles, and behavioral records are combined to predict lung cancer. [36]. In these situations, EWC offers a complementary approach, preserving important parameters from previous tasks by penalizing updates to critical weights. When integrated with replay-phase strategies, EWC enhances the model’s ability to adapt to new data while maintaining prior knowledge, ensuring more robust and reliable performance. For instance, imagine a hospital managing the care of a lung cancer patient. Integrating TNM stages, imaging data, and behavioral attributes such as smoking history into a predictive model could help clinicians project the patient’s five-year survival probability, enabling timely interventions and tailored care. Without robust hybrid CL strategies, the model’s predictions might falter as it struggles to balance new and existing knowledge. By combining replay-based methods with EWC, these challenges can be mitigated, advancing the field of survival prediction and ensuring predictive models remain reliable, adaptable, and effective in modern healthcare environments.

Another challenge is accurate feature extraction from medical imaging is critical for lung cancer survival prediction, as CT and PET scans provide valuable insights into tumor size, texture, and spread. Although, traditional methods, particularly those relying on CNNs, face significant limitations. CNNs are quite good at seeing patterns in pictures, but they frequently have trouble capturing microscopic or subtle features like tiny cancers or ground-glass opacities, which are crucial in lung cancer diagnosis and prognosis. The challenge arises because these models are typically pretrained on datasets like ImageNet, which do not capture the complex, specialized features found in medical imaging. This makes them less effective for medical applications, where the data has unique characteristics that require tailored models. Additionally, CNNs tend to focus primarily on local features, resulting in the loss of adjacent pixel and vertex information when images are resized or processed. This can hinder the detection of critical tumor features embedded in the broader anatomical context. For instance, as illustrated in Fig 1, a comparison of CNN models such as MobileNetV4, ResNet-50, VGG19, and EfficientNetV4 reveals suboptimal performance in capturing fine-grained medical imaging features between ImageNet. Feature visualization maps for these models demonstrate their dominant focus on high-level patterns rather than the intricate details needed for precise lung cancer analysis.

**Fig 1.**
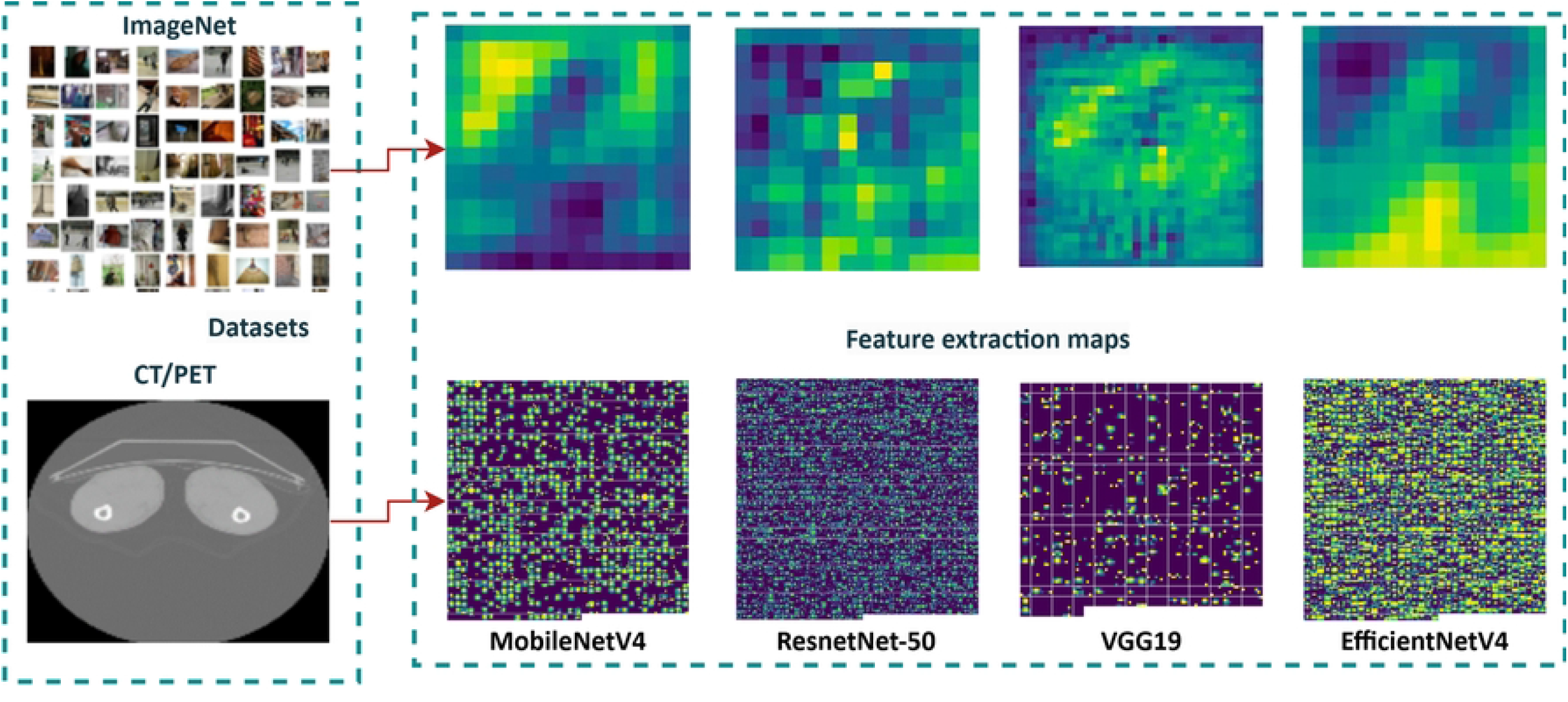
Comparison of feature extraction performance across CNN models (MobileNetV4, ResNet-50, VGG19, and EfficientNetV4) trained on ImageNet versus a medical imaging dataset. The visualization maps highlight the differences in feature focus, with ImageNet-trained models demonstrating limited sensitivity to fine-grained details.

To overcome these limitations, advanced techniques like the SwinT offer a promising alternative. Unlike Vision Transformers (ViT), which employ global attention across the entire image, the SwinT introduces Shifted Window Attention to partition images into non-overlapping windows and apply self-attention locally within these regions. This localized attention mechanism allows the SwinT to efficiently capture fine details, such as tiny tumors, while also preserving the ability to learn long-range dependencies across the image. This architectural advantage is particularly relevant for medical imaging tasks, as it enables the detection of small tumor features within a larger anatomical structure critical for accurate staging and treatment planning in lung cancer.

Furthermore, the SwinT’s ability to adapt to diverse medical imaging datasets ensures that models are not limited by biases inherent in pretrained datasets like ImageNet. By incorporating such cutting-edge techniques, this research aims to enhance imaging feature extraction and ultimately improve lung cancer survival prediction in real-world clinical settings including CL strategies.

## 4 Method

This study introduces a novel hybrid CL strategy that combines EWC and Replay-based methods within a multimodal network, enabling incremental updates of model parameters and refinement of survival predictions for lung cancer patients. The framework integrates both previously trained knowledge and newly acquired CT, PET, clinical, and DNA data. Our approach employs an enhanced feature extraction mechanism based on the SwinT, which effectively addresses the limitations of conventional CNN models pre-trained on datasets like ImageNet, which often fail to capture critical features, such as ground details and multiple tumor instances in CT and PET scans. Additionally, we leverage permutation-based XLNet techniques to learn contrastive patterns in DNA data, mitigating the challenges posed by the limited size of DNA datasets within the context of large-scale multimodal data. Clinical data are processed through an FCN network for sequential learning. To achieve adaptive and accurate survival prediction, we integrate clinical, CT, PET, and DNA data using a cross-attention fusion mechanism, which is further complemented by FCN through CoxPH modeling and a robust CL framework. The methodological overview of the proposed framework is illustrated in Fig 2.

**Fig 2.**
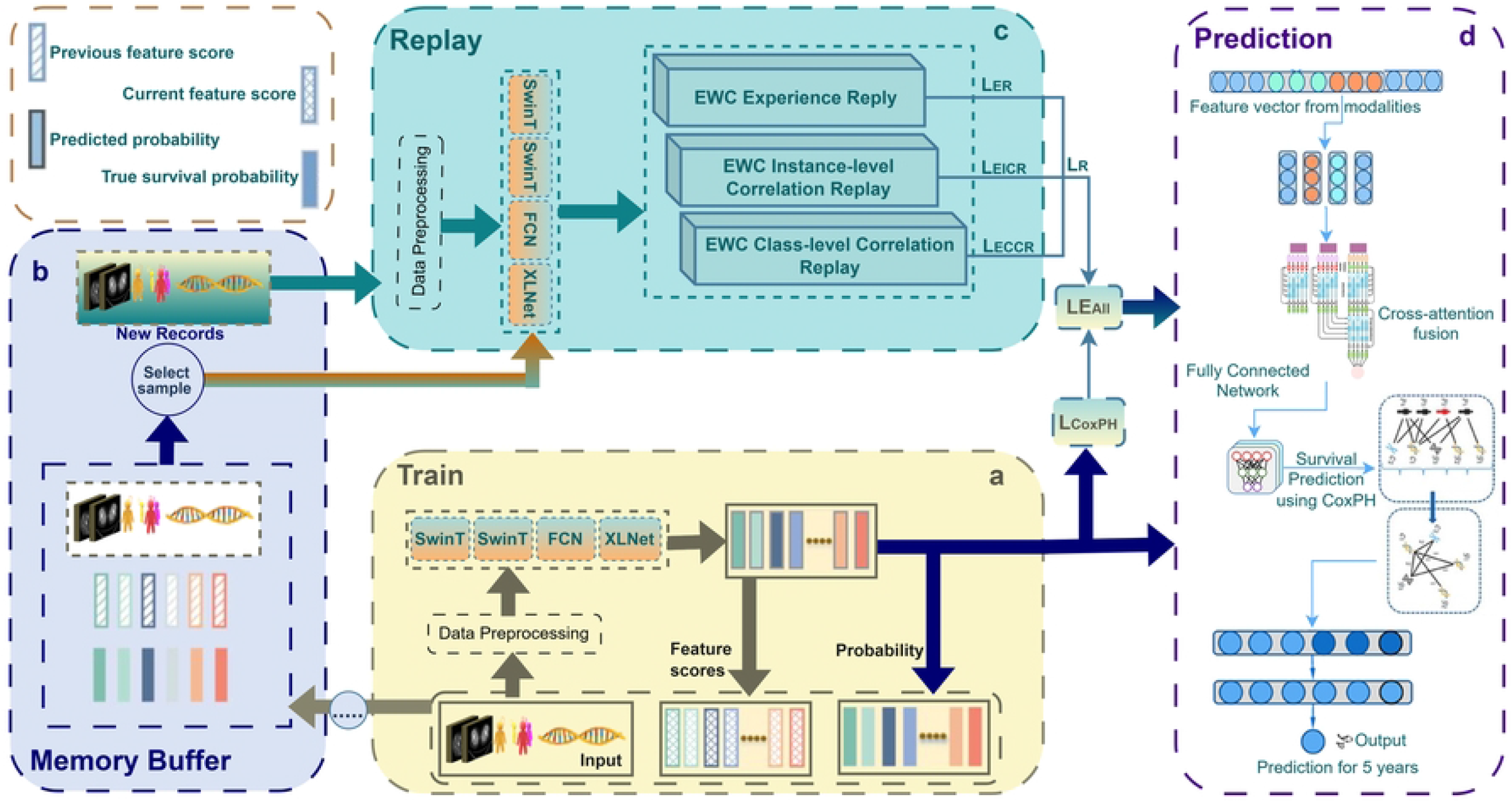
Overview of the proposed method. The framework is designed to address catastrophic forgetting in CL through a four-phase structure: training, replay, prediction, and memory buffer. During the training phase, the model sequentially learns from the current task data, organized into mini-batches. Concurrently, previously learned data stored in the memory buffer is retrieved and replayed using three hybrid EWC-based replay modules Experience, Instance-Level, and Class-Level Correlation replay which mitigate forgetting by locking important weights, applying penalties, and preserving inter-instance and inter-class relationships. Preprocessing operations are applied to both current and replayed data to enhance diversity and ensure compatibility. After the replay phase, the memory buffer is updated with representative samples from the current task, selected based on the CL strategy to maintain a compact yet informative buffer. The updated model then transitions to the prediction phase, where it estimates lung cancer survival probabilities. The legend in the upper-right corner provides additional visual guidance.

### Hybrid Incremental Learning Process

The process begins by training the base model using clinical, CT, PET, and DNA data. Clinical and imaging data are processed through FCN, SwinT, and XLNet, with a cross-attention mechanism integrating all modalities into the CoxPH framework for seamless prediction. During this phase, a replay memory (RM) buffer is initialized to store previously learned data, ensuring that valuable knowledge is preserved. As new data becomes available, it is preprocessed and used to update the memory buffer, where both old data (from previous training) and new data (from the most recent batch) are stored. The model retrains incrementally using data from the memory buffer, which includes both old and new data, to prevent catastrophic forgetting. The loss function during incremental training consists of two key components: the standard loss, which is based on the new data, and the EWC loss, which penalizes significant changes in the model’s weights to preserve previously learned knowledge. The total loss is calculated as:

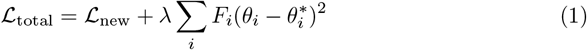

Where:

- *ℒ*_new_ is the standard loss for the new data.
- *λ* is a hyperparameter controlling the strength of the EWC penalty.
- *F*_*i*_ is the Fisher information for the *i*-th model parameter, which reflects the importance of that parameter in retaining knowledge.
- *θ*_*i*_ is the current value of the parameter, and 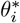 is the value from the original model.

In addition to EWC, techniques such as ER, EICR, and ECCR are integrated into the training process. The ER module aids in replaying representative samples from the memory buffer, mitigating the interference from new data by ensuring that important past information is not lost. The EICR module preserves fine-grained feature patterns by constructing a correlation matrix that captures inter-instance relationships, helping maintain structural information critical to individual data points. The ECCR module consolidates global knowledge by preserving inter-class relationships, achieved through balanced sampling and a triplet loss function that ensures clear boundaries within classes. FIM plays a crucial role in EWC by determining the significance of each parameter’s adjustment. It does this by calculating the second-order derivatives of the loss function with respect to each parameter, guiding the penalty for weight changes during incremental learning. This hybrid CL framework ensures that the model adapts to new data while retaining knowledge from previous tasks. At the conclusion of this process, the CoxPH model is used to predict the 5-year survival probability for each patient. This process is depicted in Fig 3, and a detailed explanation of each hybrid technique is provided in Fig 4.

**Fig 3.**
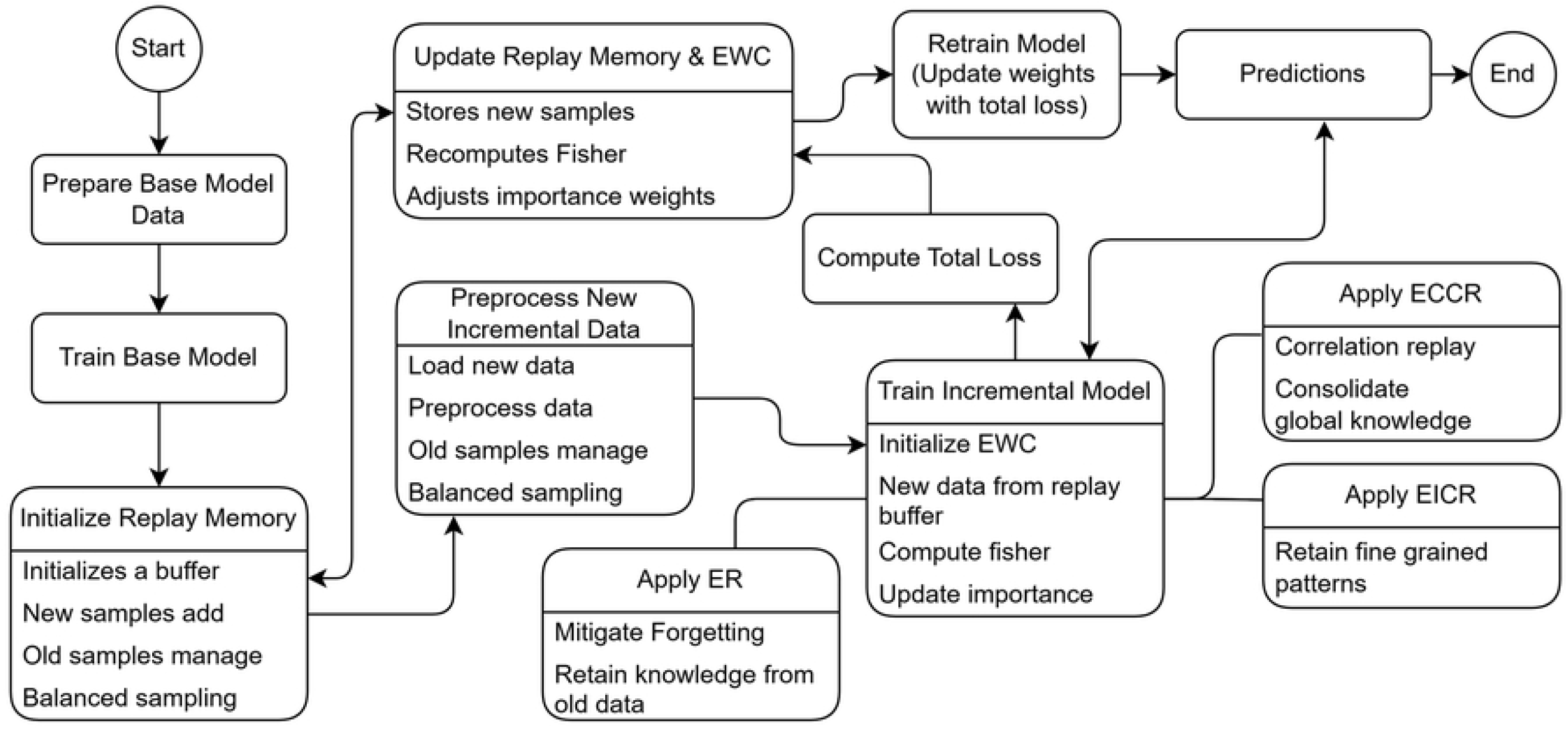
Overview of the Hybrid continual learning flowchart for incremental survival prediction.

**Fig 4.**
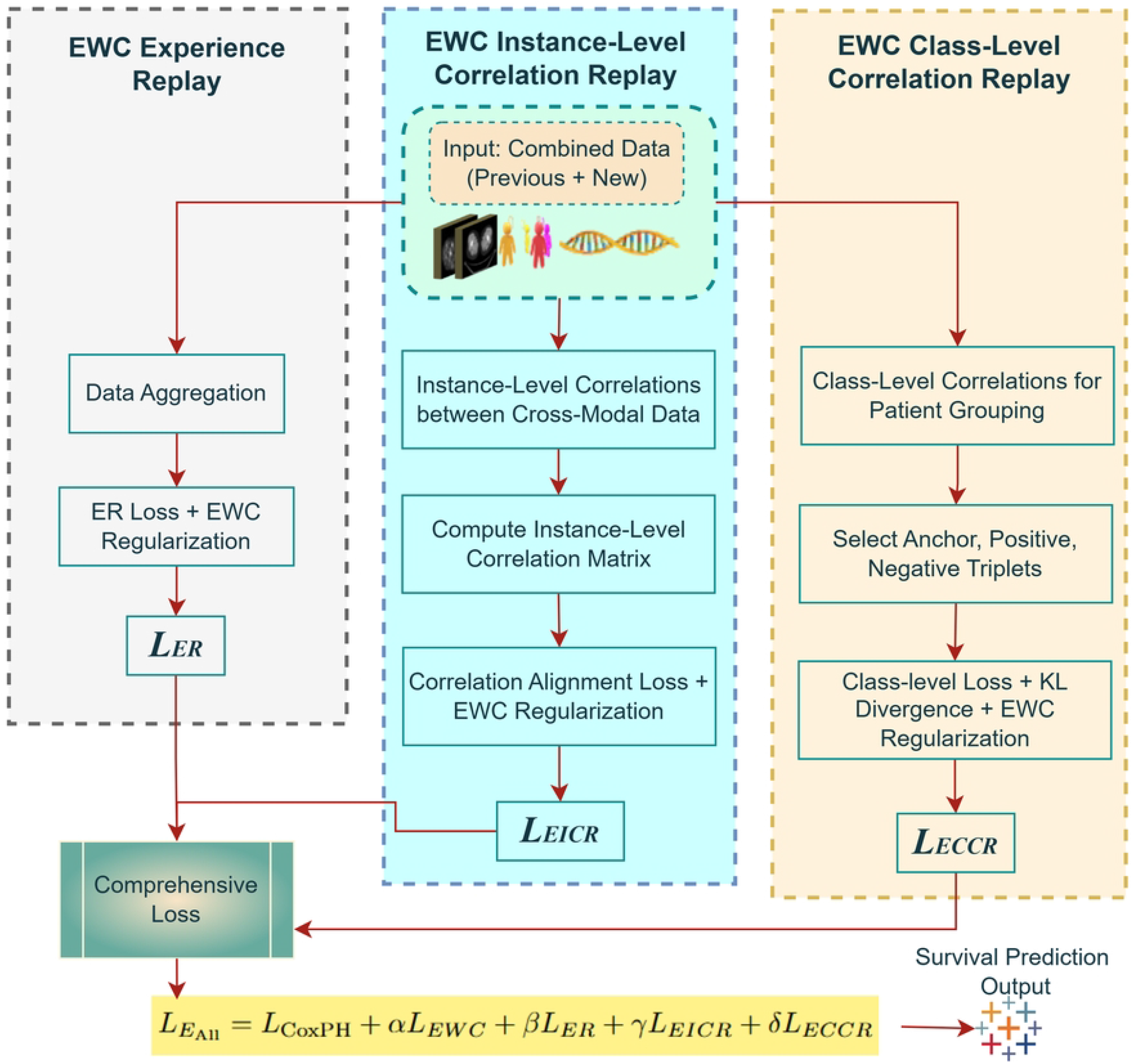
Visualization of the three types of EWC-based continual learning modules. The figure illustrates the ER Module, where past data is interleaved with new data during training to preserve knowledge. The EICR Module focuses on maintaining correlations at the instance level, ensuring model consistency for individual data samples. Lastly, the ECCR Module preserves class-level relationships by ensuring that class-specific knowledge is retained. These modules are designed to mitigate catastrophic forgetting, facilitating the continuous learning process across multiple training sessions.

### Continual Learning Approaches

In lifelong learning, a critical challenge lies in addressing catastrophic forgetting, where a model loses essential knowledge about previously learned tasks as it incorporates new data. This issue becomes particularly significant in scenarios requiring continual data integration over time, as the influx of new data risks exceeding the capacity of the memory buffer, which is designed to store only a limited subset of representative samples from prior tasks. For lung cancer survival prediction, where CL strategies remain largely unexplored, effectively managing memory constraints while ensuring adaptability to evolving data is vital. This requires innovative methods to maintain a balance between retaining past knowledge and learning from new patient data without compromising prediction accuracy. In our approach, we proposed a hybrid CL framework to tackle these challenges by combining replay-based mechanisms with EWC. Specifically, we integrate three types of hybrid techniques: ER, EICR, and ECCR. These methods aim to balance learning from new data while preserving essential knowledge from previous tasks by applying selective penalties on parameter updates and leveraging correlations among instances and classes. When new patient data arrives, it is first preprocessed and added to the memory buffer alongside representative samples from previously encountered data. During the retraining phase, the model jointly learns from this combined dataset, ensuring that the replayed data prevents catastrophic forgetting. The memory buffer selectively retains data based on importance, and penalties are applied to sensitive model parameters to protect previously learned features. This replay-based strategy enables the model to adapt to new data while maintaining performance on prior data.

Finally, during the prediction phase, the updated model predicts survival probabilities using the fused information from multiple modalities (clinical, DNA, CT, and PET). This comprehensive workflow, illustrated in Fig 2, shows the interplay of the memory buffer, replay mechanisms, and prediction phase. Furthermore, Fig 4 details the inner workings of the three hybrid CL techniques, which we discuss in the subsequent sections.

### EWC Experience Replay Module

In this module, we applied a hybrid approach combining EWC and replay strategies to address the challenges of CL in our framework. Specifically, we interleaved past patient data stored in a memory buffer with newly acquired data during each training batch. This approach enabled our model to retain prior knowledge while simultaneously integrating novel information, effectively mitigating catastrophic forgetting, as visualized in Fig 4. By revisiting historical instances alongside new data, we preserved essential patterns from prior tasks while adapting to new data. To implement this, we used experience replay to retrieve representative samples from the memory buffer and combined them with the current task data in each batch [13]. This ensured consistency in predictions across tasks. Additionally, we applied EWC regularization to safeguard model parameters critical to previously learned tasks. Together, these techniques reinforced both instance-level retention and parameter stability. The hybrid loss function we designed combines the ER consistency objective with EWC regularization as follows:

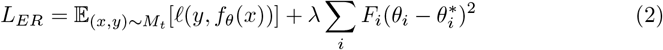

- The **first term** 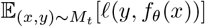 was used to calculate the cross-entropy loss for predictions *f*_*θ*_(*x*) based on past data samples (*x, y*). This ensured the retention of instance-level information from prior tasks during sequential training.
- The **second term** 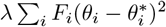 incorporated the EWC regularization penalty, where we used the FIM *F*_*i*_ to quantify parameter importance for previous tasks. By penalizing updates to critical parameters *θ*_*i*_, deviations from their original values 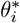 were minimized, preserving task-specific knowledge.

To compute *F*_*i*_, we precomputed the FIM at the end of each task and stored it for subsequent training phases [37]. The batch construction dynamically mixed newly acquired and replayed data, ensuring consistent representation across all training sessions. By designing this balanced loss and training strategy, we observed a significant reduction in performance degradation during task transitions, demonstrating the efficacy of this module in retaining both local and global feature patterns. Our ER module successfully enabled the model to learn continuously while maintaining robust survival prediction capabilities for lung cancer patients. This hybrid approach ensured the integration of historical and new knowledge, addressing the complexities of multimodal datasets [12, 30, 38].

### EWC Instance-Level Correlation Replay Module

We implemented the EICR module to maintain and enhance unique cross-modal relationships at the instance level, particularly those between diverse modalities such as CT, PET, clinical data, and DNA features. By integrating EWC regularization with instance-level replay, this module ensures the retention of critical inter-instance information during training, effectively mitigating catastrophic forgetting.

For this purpose, we constructed a correlation matrix *C* ∈ ℝ^*n×n*^ to capture both individual instance information and their relationships across modalities. Each instance *f*_*i*_ in the feature representation set *F* =*{ f*_1_, *f*_2_, …, *f*_*n*_*}* was processed using a high-dimensional correlation function *φ* to compute pairwise correlations:

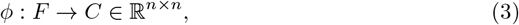

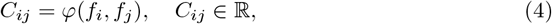

where *φ* represents the relationship between feature embeddings *f*_*i*_ and *f*_*j*_. To capture complex relationships inherent in multimodal data, we applied a high-order Taylor expansion of the Gaussian radial basis function (RBF) as follows [39]:

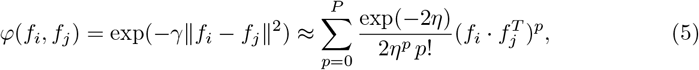

where *η* is a tunable parameter controlling the smoothness of correlations, and *p* defines the order of the expansion. This enhanced function allows the module to capture intricate dependencies among modalities, such as the interplay between DNA mutation patterns and tumor features in CT or PET scans, as well as time-series trends in clinical data.

To mitigate forgetting, we aligned correlation structures between prior training states and current data representations. The following hybrid loss function was employed to optimize this alignment:

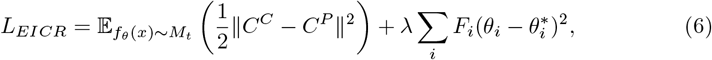

where *C*^*P*^ and *C*^*C*^ denote the correlation matrices from prior and current training states, respectively. The first term measures the frobenius norm of the difference between the matrices, ensuring that instance-level relationships are preserved across tasks [40]. The second term incorporates EWC regularization, where *λ* scales the importance of regularization, *F*_*i*_ represents the FIM [37], and 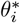 denotes the initial values of parameters from earlier tasks. During implementation, we stored correlation matrices for each modality in the memory buffer and dynamically updated them during training. For example, DNA data correlations were computed to emphasize mutation patterns, while CT and PET features highlighted tumor intensity or spatial relationships. Clinical data, processed through an FCN network, captured sequential trends, such as progression markers over time. At the end, this module ensures the preservation of critical cross-modality relationships, such as correlations between specific mutations and tumor growth, during task transitions. As illustrated in Fig 4, the process includes correlation matrix computation, feature embedding alignment, and EWC regularization. By leveraging the EICR module, our framework adapts effectively to new datasets while retaining essential information for accurate survival predictions in lung cancer patients.

### EWC Class-Level Correlation Replay Module

In this stage of our experiments, we developed the ECCR module to preserve class-specific knowledge across different patient groups, ensuring consistent survival prediction for categories such as early-stage and late-stage cancer patients. To achieve this, we combined EWC with a triplet-based correlation mechanism tailored to our multimodal dataset, which includes CT, PET, clinical, and DNA data. This approach allowed us to maintain class-level distinctions while learning new data without catastrophic forgetting [30]. We implemented a triplet mechanism where, for each anchor *x*_*i*_ (e.g., a patient with early-stage cancer), we selected a positive sample *x*_*j*_ (another early-stage patient) and a negative sample *x*_*k*_ (a late-stage patient), as shown in Fig 4. This setup allowed us to retain both intra-class similarities and inter-class differences. For example, CT and PET tumor intensity patterns were used as anchor-positive pairs within the same class, while DNA mutations or clinical time-series data helped distinguish anchor-negative pairs. The triplet loss was computed as follows:

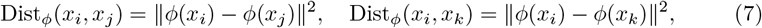

where *ϕ*(*x*) represents the fused feature embedding obtained through cross-attention layers processing data from all modalities. To ensure meaningful class-level separation, we applied a semi-hard triplet selection strategy, where negative samples *x*_*k*_ were chosen such that:

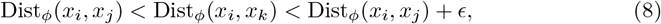

with *ϵ* defining the margin. For example, genomic variations were used to find samples that were similar but not identical within the same class. In our implementation, the triplet mechanism was complemented with a probabilistic alignment strategy. For each triplet, we computed the class-level probability distribution as:

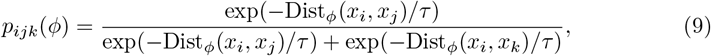

where *τ* is the temperature parameter. This probabilistic distribution helped us measure how well class separability was maintained across training updates. To align prior and current training phases, we minimized the Kullback-Leibler divergence between Bernoulli distributions derived from past (*P*^*P*^) and current (*P*^*C*^) class-level probabilities [41]:

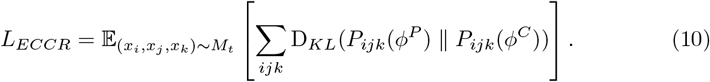

This loss ensured the preservation of key class-level patterns during training. In our framework, we combined the ECCR loss with EWC to protect critical weights from past tasks:

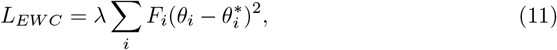

where *θ*_*i*_ and 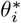 represent the current and prior weights, respectively, and *F*_*i*_ denotes fisher information for each parameter.

Finally, the combined objective function for training included the survival prediction loss *L*_CoxPH_, along with replay and regularization terms:

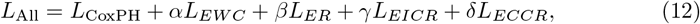

where *α, β, γ*, and *δ* are hyperparameters that balance the contributions of each term. Specifically: - *L*_*ER*_ represents the ER loss for direct replay of past data. - *L*_*EICR*_ is the instance-level correlation replay loss. - *L*_*ECCR*_ is the class-level correlation replay loss, as described above in the hybrid loss function. Our experimental setup involved extracting multimodal features from CT and PET images using a SwinT, which captured spatial and intensity-based tumor characteristics. DNA data, including mutation metrics, was processed using XLNet to learn latent genomic features, while clinical data, such as lab records and vital signs, was modeled using FCN networks to capture temporal patterns. These features were fused using a cross-attention mechanism to create a unified embedding. The embeddings were then processed through the ECCR, ER, and EICR modules, where the ECCR ensured class-level correlation learning by maintaining intra-class similarity and inter-class dissimilarity. The output of these modules was passed to the FCN, which further processed the learned features. Finally, during the prediction phase, the model used the CoxPH loss function (*L*_CoxPH_) to compute survival probabilities, with guidance from the hybrid loss function. The integrated mechanisms of ER, EICR, and ECCR played critical roles in preserving important patterns from previous tasks while enabling the incorporation of new patients information [42]. By preserving class-level relationships and aligning past and current knowledge through EWC-based regularization [30], the model ensures that it does not forget previously learned survival patterns while adapting to new data. This process enables the model to predict the 5-year survival probability for each patient, with stable and accurate predictions over time.

### Clinical Modality Processing with FCN

We utilized 16 clinical features, including demographic, clinical staging, and behavioral factors, for survival prediction. These features were preprocessed using standard tabular techniques. In our process, FCN is designed to generate a token embedding (token dim = 64) that aligns with other modalities. The 16 clinical features (input dim = 16) pass through the FCN, which progressively reduces dimensionality across layers with 512, 256, 128, and finally 64 neurons. Each layer incorporates batch normalization for stability, ReLU for non-linearity, and Dropout (0.3) to minimize overfitting. This hierarchical structure captures increasingly abstract features, ensuring efficient feature representation. The final 64-dimensional token was chosen empirically to balance computational efficiency and performance. Its compatibility with other modalities, which also output 64-dimensional tokens, we ensured seamless integration during multimodal fusion. Mathematically, given the input *X* ∈ *ℝ*^*N×D*^, where *N* is the batch size (32 samples per batch) and *D* = 16 is the feature dimension, the FCN reduces *D* through hierarchical transformations as follows [43]:

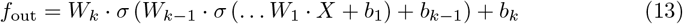

Where: *W*_*i*_, *b*_*i*_ represent the weights and biases of the *i*-th layer, *σ* is the ReLU activation function, *f*_out_ ∈ ℝ^*N×*64^ is the final token embedding.

These embedding tokens effectively represent the clinical features while maintaining consistency with the token outputs from other modalities. These tokenized clinical features are subsequently passed to the cross-attention fusion block, where they are integrated with the tokenized outputs from CT, PET, and DNA data. This cross-modality integration enhances the survival prediction model by combining temporal insights from the clinical data with spatial features from imaging modalities and genomic data, ultimately improving the survival prediction process. The stage of FCN in the framework is illustrated in Fig 2.

### DNA Modality Processing with XLNet

In our multimodal framework, we used XLNet to process DNA features, such as the counts of SNVs, HOMO, and HETE variants, to extract meaningful patterns for lung cancer prediction. To enable this, we configure XLNet with specific parameters to capture complex dependencies in the genomic data. The model is configured with a vocabulary size based on the number of unique tokens from the data. We set the number of layers to 6, with 4 attention heads in each layer. Each attention head has a dimensionality of 8, and the inner layer dimension (for the feed-forward layer) is set to 32, which ensures the model can capture intricate relationships between DNA features. The dimensionality of the model we set to 32, which directly determines the size of the embedding space for each token. We employed permutation-based autoregressive modeling, where the model predicts each token *x*_*t*_ conditioned on the previous tokens in a randomly permuted sequence for learning the pattern of data. The task is formulated as:

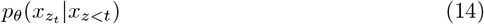

Here, *p*_*θ*_ denotes the probability of predicting the token 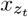 at position *z*_*t*_, given the preceding tokens *x*_*z<t*_. This permutation-based approach allows XLNet to model bidirectional context efficiently by learning from all possible orderings of the sequence. By leveraging a multi-head attention mechanism to capture different aspects of relationships between tokens at various positions. The attention mechanism in each layer represented as:

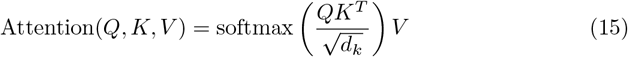

where *Q* is the query matrix, *K* is the key matrix, *V* is the value matrix, and *d*_*k*_ is the dimension of the key vectors. In our configuration, we use 4 attention heads, with each head having a dimensionality of 8, allowing the model to capture diverse relationships between the features, such as SNVs, HOMO, and HETE variants. The multi-head mechanism enables the model to learn complex dependencies at multiple levels of granularity. Through the self-attention mechanism, it learns latent patterns in the DNA data by attending to different parts of the input sequence. The self-attention layer updates the input embeddings by calculating:

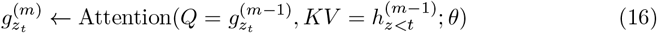

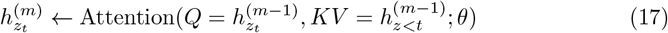

We utilized the iterative update process for both the query stream and content stream, which progressively refine the embeddings at each layer. The learned latent patterns are crucial for understanding the relationships between different genomic features. After processing the DNA data through multiple XLNet layers, the final output embeddings *h*^(*M*)^ are projected into a 64-dimensional space through a linear transformation. This transformation is performed by a fully connected layer, as presented:

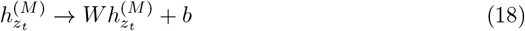

where the output embeddings are multiplied by a learned weight matrix 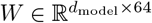, followed by an optional bias term *b* ∈ ℝ^64^. This ensures that the final embedding dimension matches the required token size of 64. The final tokens are passed into the cross-attention fusion block, where they are merged with embeddings from other modalities, such as CT/PET and clinical, for multimodal integration, as shown in Fig 5. This fusion allows the model to leverage complementary information across modalities to improve prediction accuracy.

**Fig 5.**
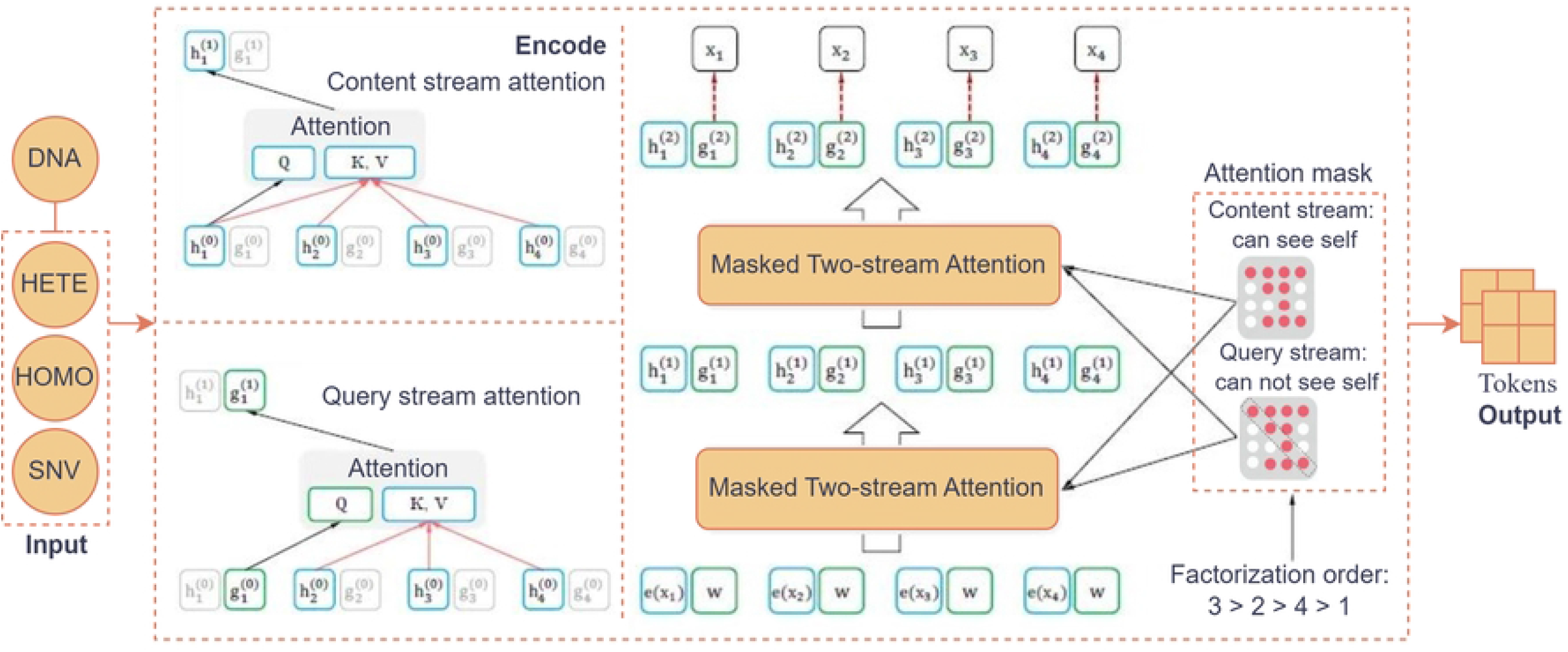
XLNet architecture for DNA data. This figure illustrates the model applied to DNA data, including SNV, HOMO, and HETE counts. Then it tokenizes the input sequence and uses permutation-based autoregressive training to capture dependencies among features. A Two-Stream Self-Attention mechanism, consisting of a Query Stream for prediction and a Content Stream for context encoding, is applied to generate contextual token representations. These enriched token embeddings are then fused with other modality embeddings for prediction analysis.

### CT and PET Modality Processing with SwinT

We implemented the SwinT model as an efficient approach for processing high-resolution medical images, such as CT and PET scans. It addresses the limitations of ViT, particularly in terms of computational complexity, while offering improved performance in classifying fine-grained object details compared to traditional CNNs. This makes SwinT particularly well-suited for medical imaging tasks, where high accuracy in identifying complex structures, such as ground objects and tumors in CT/PET scans, is crucial [44]. We begin the input shape and preprocessing of CT/PET image with a shape of [1, 160, 128, 128], where:1 represents the batch size, 160 is the depth (number of slices), and 128 *×* 128 are the spatial dimensions of each slice. The patch embedding of The image is divided into non-overlapping patches of 4 *×* 4 pixels, resulting in tokens of shape [*N, C*], where *N* = 160 *×* 128 *×* 128*/*4^2^ = 4096 patches and *C* is the initial feature dimension. Each patch is flattened and projected to a higher-dimensional space via a linear embedding layer. In the Window Multi-Head Self-Attention (W-MSA) operation we computes attention within fixed windows, reducing complexity. Attention is calculated within windows of size *M × M*, reducing computational complexity to linear with respect to the number of patches:

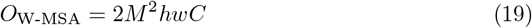

After each W-MSA block, in Shifted Window Multi-Head Self-Attention (SW-MSA) windows are shifted to capture cross-window dependencies. This introduces quadratic complexity:

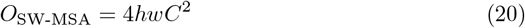

These steps enhance both local and global context modeling. After processing through several SwinT blocks, the output tokens are passed through a Multi-Layer Perceptron (MLP) to refine features, followed by Layer Normalization for dimensionality reduction. Finally, token embeddings 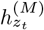 are projected into a 64-dimensional space using a fully connected layer. This is performed by multiplying the embeddings with a learned weight matrix 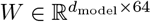, where *d*_model_ is the feature dimension before projection, followed by an optional bias term *b* ∈ ℝ^64^. This ensures the output token size is reduced to 64, making it compatible for integration into downstream tasks like survival prediction:

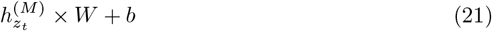

After tokenization and projection, the features from both CT and PET modalities are passed into a cross-attention fusion block, which combines the features with clinical, imaging, and mutation data. where the features from both modalities are merged using attention mechanisms. This fusion ensures that the combined representations are more informative and relevant for the downstream survival prediction task. The methodological overview of this process is illustrated in Fig 6.

**Fig 6.**
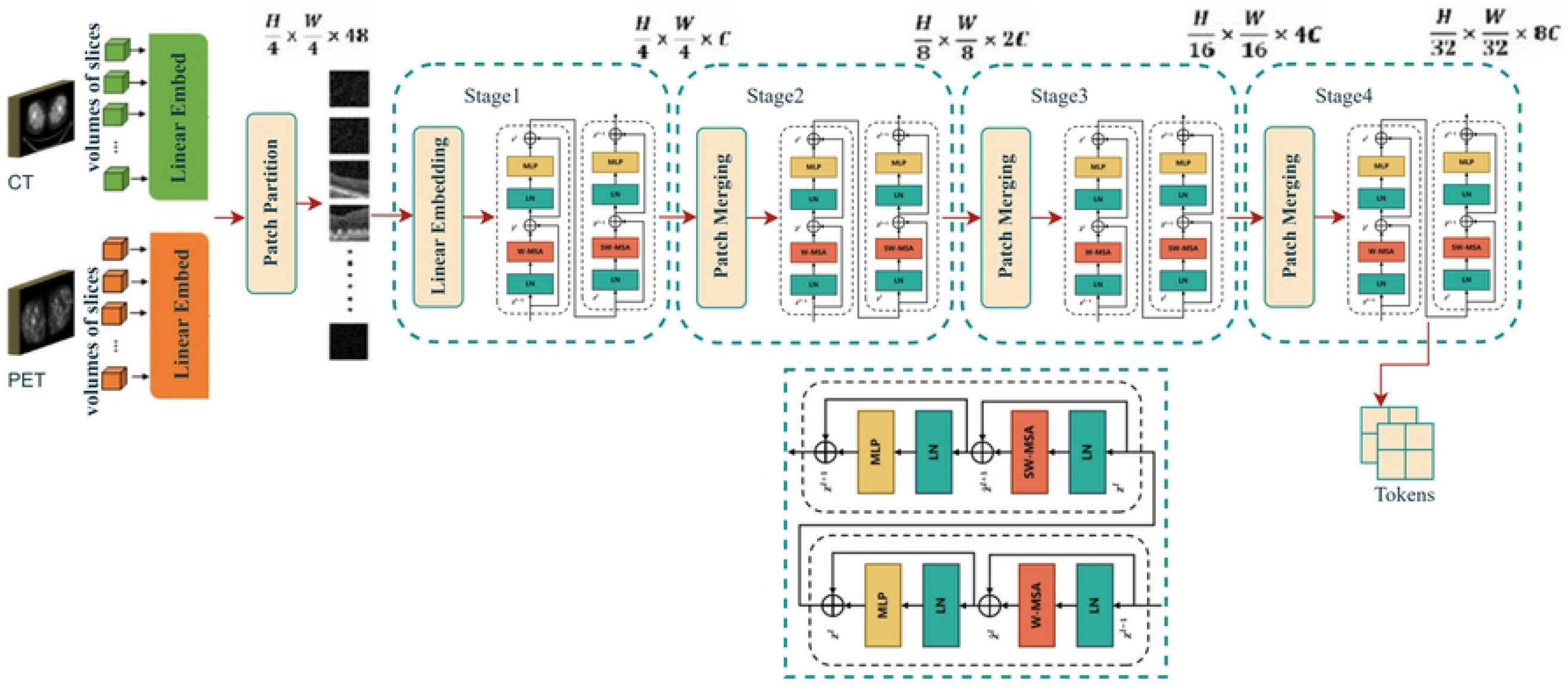
The overall architecture of the Swin Transformer adapted for CT and PET scan analysis. The architecture consists of two successive Swin Transformer blocks, each designed to process image patches from CT and PET scans, enabling the extraction of multi-scale feature representations. The hierarchical structure enhances the model’s efficiency in handling high-resolution medical images by reducing computational complexity. The figure also illustrates the shifted window strategy for computing self-attention, which helps the model capture long-range dependencies and fine-grained details, such as tumors and ground objects, across different regions of the dicom slices.

### Cross-Attention Fusion Block

In our survival prediction framework, inspired by recent works such as [45], the cross-attention block is employed to integrate clinical, DNA, CT, and PET data by capturing dependencies both within and across these modalities. The data from each modality is first formatted into a consistent *L × D* structure, where *L* is the number of tokens or features, and *D* represents the feature dimension. Specifically, the CT and PET images are split into patches, tokenized, and flattened, resulting in a sequence of tokens (of length *L*_img_), where each token represents a patch encoded into *D*-dimensional space. For clinical and DNA tabular data, individual attributes or features are embedded directly into *L*_tab_ *× D*, ensuring that all modalities share a common format, making cross-attention feasible.

The core of the cross-attention mechanism utilizes this unified structure to link information across modalities. The non-local operation within the cross-attention block is defined as:

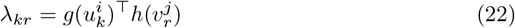

where 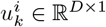 represents the *k*-th feature embedding of modality *i*, and 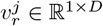 represents the *r*-th feature embedding of modality *j*, with *g* and *h* as learned transformations that optimize compatibility between the features of the modalities. Here, *i* ≠ *j* ensures cross-modal interactions, such as between CT data and clinical attributes or PET data and DNA features.

To compute the attention map *P*^(*i*,*j*)^ that captures the relevance between tokens from different modalities, we calculate:

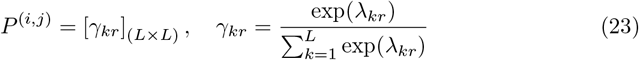

Each element *γ*_*kr*_ represents the attention score between a pair of tokens from different modalities, capturing the degree of relevance between them.

Once the attention weights are calculated, these cross-modal connections are used to refine the features by aggregating relevant information across modalities. For each pair of modalities, the feature representations are updated as follows:

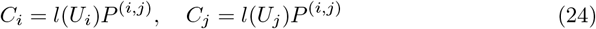

where *C*_*i*_ and *C*_*j*_ represent the fused feature representations of each modality, and *l*(*U*_*i*_) and *l*(*U*_*j*_) denote learned transformations applied to the feature matrices *U*_*i*_ and *U*_*j*_, respectively. The resulting features are adjusted by non-negative coefficients *β*_*i*_ and *β*_*j*_ to preserve their individual relevance.

Finally, the fused output of the cross-attention block is obtained by concatenating the refined features:

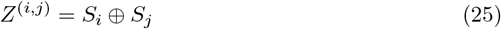

where *S*_*i*_ and *S*_*j*_ are the enhanced, modality-specific feature maps, and ⊕ represents the concatenation operation. This cross-attention-based fusion enables our model to utilize the comprehensive, interrelated feature representations from both image and tabular modalities. The output of the cross-attention, represented by the fused features *Z*^(*i*,*j*)^, is passed to the FCN phase for further processing. The FCN layer refines these multimodal features and extracts relevant patterns for survival prediction. The mechanism of cross-attention fusion is displayed in Fig 7.

**Fig 7.**
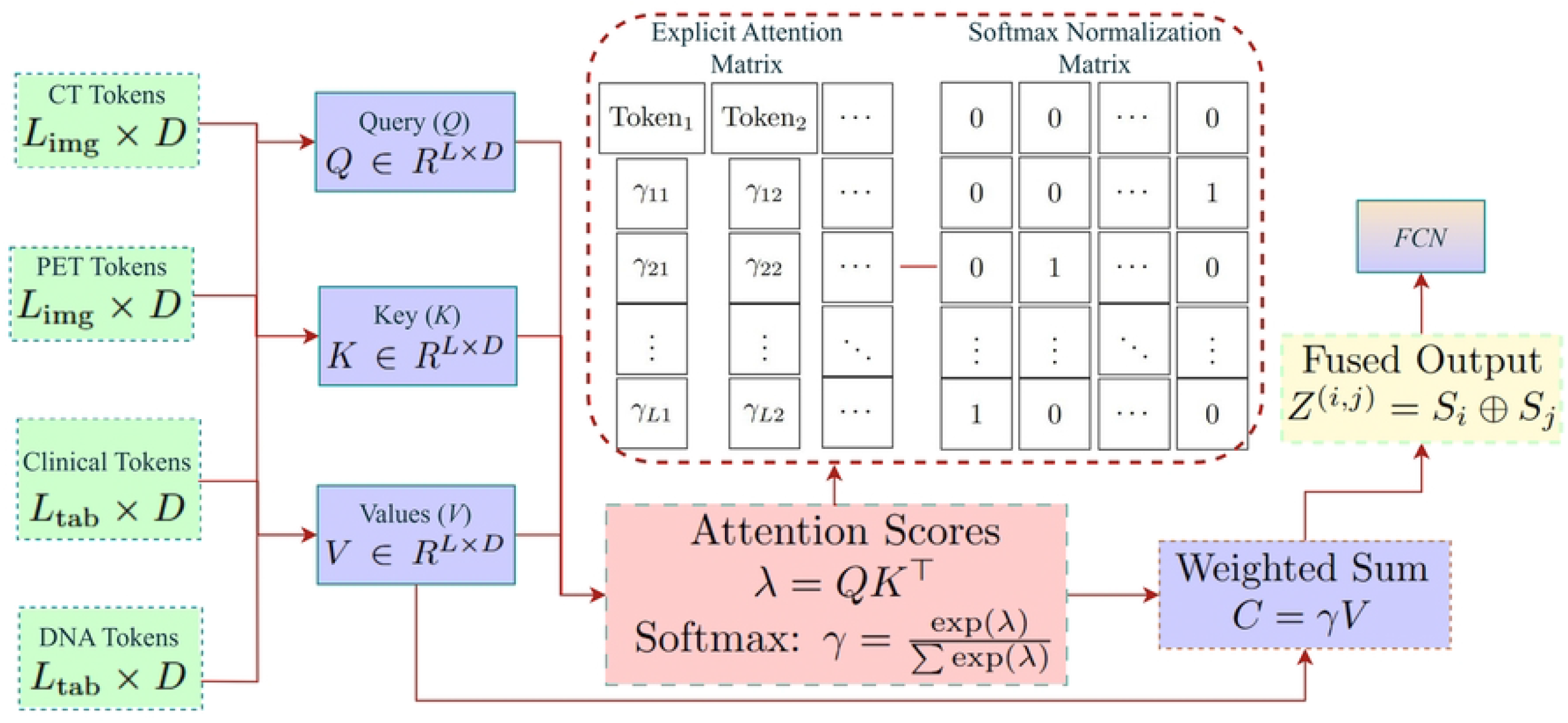
Illustration of the cross-attention mechanism for multimodal data fusion. clinical, DNA, CT, and PET data are embedded into a unified feature space. Cross-modal attention is computed between feature pairs, updating and refining modality-specific representations, which are then concatenated for further processing by the FCN in the survival prediction model.

### Fully Connected Network Phase

In this FCN phase, we refines multimodal feature embeddings from the cross-attention module into a predictive representation for survival analysis. The FCN comprises three dense layers, each progressively reducing dimensionality and capturing intricate relationships within the integrated data. Between these layers, dropout layers with a rate of 0.3 are applied to mitigate overfitting and enhance the model’s robustness on unseen data. The final dense layer produces a feature vector using linear activation, which is sent to the CoxPH model for 5-year survival prediction. This structured pipeline, illustrated under the **prediction** process in Fig 2, ensures comprehensive risk stratification by leveraging multimodal data. The approach aligns with the design principles of robust multimodal deep learning frameworks such as those discussed in [46, 47].

### Cox Proportional-Hazard Workflow

We implemented the CoxPH model, a cornerstone of survival analysis, to process the feature vector derived from the FCN and estimate 5-year survival probabilities. The hazard function is expressed as:

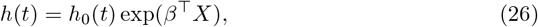

where *h*_0_(*t*) is the nonparametric baseline hazard, *β* is the vector of learned regression coefficients, and *X* is the FCN-generated feature vector. The survival probability *S*(*t*) is computed as:

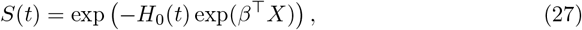

with *H*_0_(*t*), the cumulative baseline hazard, estimated using methods like the Breslow estimator. We utilized the CoxPH model semi-parametrically, as it does not require specific distributional assumptions about survival times, ensuring flexibility across diverse datasets [48, 49]. Integrated into our continual learning (CL) framework, the CoxPH model dynamically adapts to new patient data while preserving critical knowledge from prior data. We used techniques such as EWC [30] and experience replay [50] to ensure that new learning does not overwrite essential model parameters. For example, when incorporating new features, we recalibrated the baseline hazard *h*_0_(*t*) without disrupting existing regression coefficients *β*, maintaining stable survival prediction accuracy. Leveraging the PyCox library [51] (An algorithmic 14 summary is presented below.), we integrated the CoxPH model with FCN outputs to enable seamless functionality and facilitate periodic updates, enhancing the flexibility and adaptability of our survival prediction framework. This implementation combines semi-parametric modeling with dynamic learning capabilities, making it particularly effective for our multimodal survival prediction pipeline. By continuously learning from both clinical and imaging data, the framework ensures stability in previously learned parameters while maintaining consistent performance over time. As illustrated in Fig 2, under the **prediction** section, our continual learning approach enables accurate estimation of 5-year survival probabilities while supporting robust adaptability to new data.

#### Algorithm 1 CL Workflow for Lung Cancer Survival Prediction

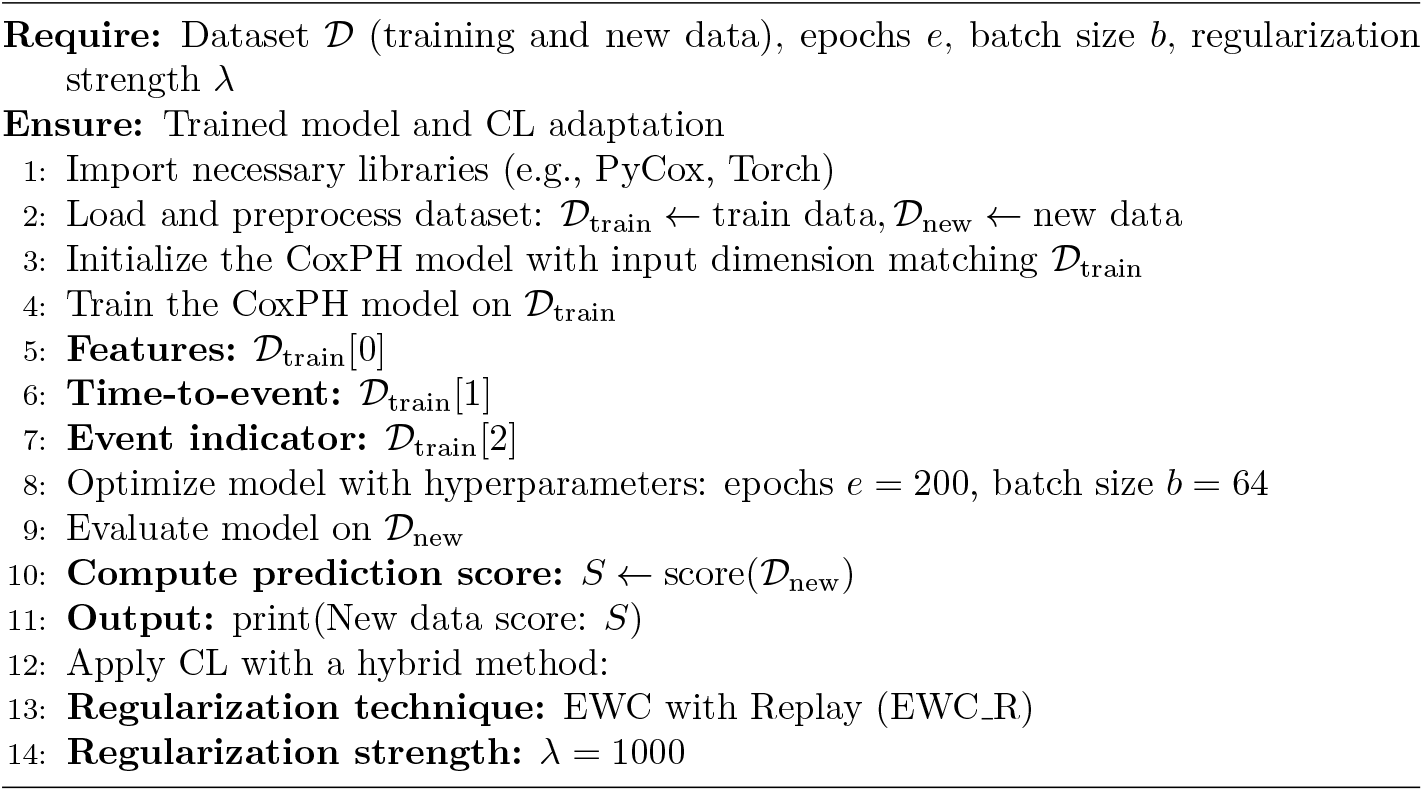

## 5 Experimental Setup

### Dataset

We accessed a comprehensive dataset from aihub [52], comprising records of 5,053 lung cancer patients. Among these, 3,770 patients had complete clinical, CT, PET, and DNA data, alongside detailed survival investigation timelines spanning up to four years. However, DNA mutation data was available for only 412 patients. To address this limitation, we utilized XLNet, a transformer-based model, to learn and encode patterns from the available DNA data, rather than directly training the model on the sparse DNA information. This pretraining step generated meaningful representations of DNA features, which were subsequently integrated into the multimodal framework, alongside clinical, CT, and PET data. For evaluation, we divided the dataset into three subsets. The primary training dataset included 3,358 patients (**D_3358P**), incorporating clinical, CT, PET, and XLNet-derived DNA embeddings. To test the model’s CL capabilities, two additional datasets were constructed to simulate incremental data updates. The first set consisted of 200 patients (**D_200P**), and the second included 212 patients (**D_212P**), these retaining equivalent real DNA mutation data alongside the other modalities. This multi-dataset strategy allowed us to effectively evaluate the model’s adaptability, ensuring it could integrate new knowledge while maintaining stability in previously learned survival patterns.

### Clinical Data Preprocessing

In the data preprocessing phase, we handled four distinct modalities: clinical data, CT and PET images, and DNA mutation data. For clinical data, we utilized 16 features, including essential variables like patient records (PatientID, Gender, Age, survival time, deadstatus event, overall stage, and clinical stages (TNM), along with attributes such as smoking status and smoking amount. Common tabular preprocessing techniques were applied, including outlier detection using the Zscore method, one-hot encoding for categorical variables, and normalization to ensure data consistency. Redundant information, such as description, histology, and FILE NAME, were dropped. The target variables were identified as survival time’ and deadstatus event which were crucial for survival prediction and then feed into the FCN.

### CT/PET Data Preprocessing

We developed a comprehensive preprocessing pipeline for CT and PET imaging data to ensure the input is standardized and optimized for downstream analysis using the SwinT model. First, we configured the target dimensions for the images, setting each slice to 128×128 pixels and the depth to 160 slices. These specifications ensured consistent input dimensions for all scans, which is crucial for effective processing by the SwinT. We loaded the DICOM files for each patient and sorted the slices based on their SliceLocation metadata to preserve the anatomical sequence. Each slice was resized to the target spatial resolution using bilinear interpolation (a 2×2 grid neighboring pixels), while the 3D volumes were padded or interpolated to match the desired depth. For scans with fewer slices, we applied padding with appropriate background values (−2000 for CT scans and 0 for PET scans) to maintain anatomical integrity. If the number of slices exceeded the target depth, we used cubic spline interpolation (cubic polynomial between data pixels points) to compress the scans, ensuring they retained their structural relevance. Finally, the processed volumes were reshaped into a uniform format of [1, 160, 128, 128], suitable for feeding into the SwinT. To enhance generalization and minimize overfitting, we applied random data augmentations to the 3D volumes. These included horizontal and vertical flips, small random rotations (±10 degrees), and the addition of Gaussian noise to simulate real-world variability in imaging data. After augmentation, the data was normalized to a [0, 1] range to ensure consistency in pixel intensity values, which aids in stabilizing the training process for the DL model. The preprocessed data was split into training, validation, and testing subsets with an 80-10-10 split. This ensured a representative distribution across all subsets, avoiding any bias in the model’s evaluation. The processed CT and PET images were then stored as files for efficient loading and reuse during training. The prepared 3D image volumes were fed into the SwinT for learning.

### DNA Data Preprocessing

The DNA data preprocessing workflow involves systematically preparing raw genomic information contained in tsv files for integration into our learning framework. This process ensures that genomic features are accurately extracted, cleaned, normalized, and formatted for downstream modeling. The input data comprises multiple tsv files, each representing a single patient’s genetic data. These files include information, which identifies the variant type, and ZYGOSITY, which describes whether a variant is homozygous (HOMO) or heterozygous (HETE). The goal of the preprocessing is to summarize these genetic variations into meaningful metrics while addressing data inconsistencies and ensuring compatibility with our lung cancer pipelines. First, we read all files and loaded them into a pandas DataFrame, where any empty lines or encoding issues are resolved. A series of cleaning operations follow, including the removal of rows with missing or invalid entries in the TYPE or ZYGOSITY. If a file is found to be empty or contains no valid data after cleaning, it is filled with nearby mean values to maintain the integrity of the dataset. Thern from the cleaned data, three critical genomic features for lung cancer are extracted for each patient: the count of single nucleotide variants (SNVs), HOMO, and HETE variants. SNVs are counted based on rows where the TYPE column contains SNV, while the counts of HOMO and HETE are derived from the ZYGOSITY. These metrics are key indicators in understanding genetic variations, particularly in cancer genomics, as they provide insights into mutation burdens and genetic heterogeneity within tumors. After feature extraction, normalization is performed to standardize the values across patients. Using min-max scaling, the raw counts of SNVs, HOMO, and HETE variants are transformed to a scale between 0 and 1. This step ensures that genomic features have comparable influence when combined with other data modalities, such as clinical and imaging data. Finally, we saved the processed and normalized data into a summary file. Finally, the data are fed into the XLNet for further processes of latent pattern study.

Throughout all preprocessing steps, patient IDs were synchronized across modalities to maintain data integrity and prevent mismatched entries. This alignment ensured consistency during training, evaluation, and CL, where new data undergoes similar preprocessing pipelines to integrate seamlessly with the existing data.

### Pre-training Setting

All experiments were conducted using a default pre-training configuration of 200 epochs unless specified otherwise. To optimize model performance, we employed the AdamW optimizer with a batch size of 32, ensuring efficient weight updates and regularization. The implementation was carried out using the PyTorch framework, leveraging its flexibility for model design and training. The computational setup included 2 NVIDIA GeForce RTX 3070 GPUs, 64 GB of RAM, and an Intel(R) i9-10900 processor, providing sufficient resources for handling complex computations and ensuring smooth training. This configuration was chosen to balance efficiency and scalability while maintaining consistent results across investigations.

### Evaluation Setting on Survival Prediction

To assess the performance of our CL-based multimodal framework, we employed widely recognized metrics, including the concordance index (C-index), which measures the predictive accuracy of survival models by evaluating the concordance between predicted and actual survival times [53], and Mean Absolute Error (MAE), which quantifies the average magnitude of errors between predicted and observed survival times [54]. For the evaluation of CL, we utilized key criteria: Baseline Performance (BP, accuracy on the initial dataset), New Data Performance (NDP, accuracy on new, unseen data), Knowledge Retention (KR, ability to retain previously learned information), and Forgetting (Fg, degree of degradation in earlier learned tasks) [28, 30]. Additionally, we compared our framework against state-of-the-art multimodal survival prediction models to highlight its effectiveness. An ablation study was conducted to analyze the contribution of each modality and the cross-attention mechanism within the CL workflow, providing deeper insights into the impact of individual components.

## 6 Results and Discussion

### Ablation studies

In the ablation study, we conducted a series of performances of the proposed CL framework for lung cancer survival prediction by incrementally modifying its components. These experiments evaluate the effectiveness of strategies such as EWC, replay mechanisms, cross-attention fusion, and the inclusion of DNA embeddings. To ensure fair comparisons, all experiments are conducted with the same hyperparameters: batch size of 32, the AdamW optimizer, learning rate of 1e-4, and weight decay of 1e-4. Evaluation metrics include C-Index, MAE (in days), KR, and Fg, as shown in Table 1. The component-wise evaluations are discussing below:

**Table 1.** Ablation Study for Lung Cancer Survival Prediction Framework.

| Datasets Used: | D_3358P, D_200P, and D_212P |  |  |  |
| --- | --- | --- | --- | --- |
| Model Variant | C-Index (↑) | MAE (Day ↓) | Knowledge Retention | Forgetting (↓) |
| HCLmNet (Proposed) | **0.84** | **140** | **0.86** | **0.08** |
| ✗ CL Mechanism | 0.78 | 180 | 0.25 | 0.83 |
| ✗ DNA Modality | 0.81 | 155 | 0.66 | 0.26 |
| Raw DNA Features → XLNet | 0.79 | 260 | 0.64 | 0.19 |
| Concatenation → Cross-Attention | 0.80 | 168 | 0.65 | 0.35 |
| CL Replay → EWC | 0.82 | 155 | 0.63 | 0.16 |
| CL EWC → Replay | 0.83 | 190 | 0.71 | 0.45 |

**Notes:** - C-Index (↑): Higher values indicate better discriminative ability. - MAE (↓): Lower values reflect higher accuracy in survival time prediction. - Knowledge Retention: Performance retention metric post-training with new data. - Forgetting: Measures performance degradation on old data.

#### HCLmNet (Proposed)

The full framework integrates a FCN for clinical, SwinT for CT/PET feature extraction, XLNet for generating DNA embeddings, cross-attention for multimodal fusion, and CoxPH for survival prediction. Combined with both EWC and replay mechanisms, this configuration achieves the best results, with a C-Index of 0.84, MAE of 140 days, KR of 0.86, and minimal Fg at 0.08. The superior performance underscores the importance of integrating all these elements for effective survival prediction.

#### Without CL Mechanism

Eliminating both EWC and replay mechanisms leaves the model susceptible to catastrophic forgetting, resulting in degraded performance. The C-Index drops to 0.78, MAE increases to 180 days, and KR decreases to 0.25, while Fg increases to 0.83. This highlights the critical role of CL in preserving knowledge during incremental updates.

#### Without DNA Modality

For checking the diversity of input data we removed the DNA modality. Then the framework processes clinical, CT and PET features only. Consequently, the C-Index falls to 0.81, MAE rises to 155 days, and KR drops significantly to 0.66, with Fg increasing to 0.26. These results emphasize the significance of DNA data in improving predictive accuracy.

#### Embedding with Raw DNA Features

Here, we replaced the XLNet embeddings with raw DNA mutation features eliminates pretrained DNA representations, leading to poorer performance. The C-Index decreases to 0.79, MAE increases sharply to 260 days, KR drops to 0.64, and Fg rises to 0.19. This demonstrates the advantages of leveraging advanced integrating for DNA data.

#### Using Concatenation Instead of Cross-Attention

In this variant we replaces cross-attention with feature concatenation, which removes the dynamic alignment and weighting of multimodal inputs. The resulting C-Index is 0.80, MAE increases to 168 days, and KR drops to 0.65, with Fg increasing significantly to 0.35. The findings underline the superiority of cross-attention for multimodal fusion.

#### CL Replay method apply Instead of EWC

In this configuration, the framework retains only the replay mechanism, relying on a memory buffer to revisit past examples during training. However, without EWC, which regularizes parameters by preserving critical weights, the model becomes susceptible to information leakage, where significant features from earlier tasks are not adequately preserved in subsequent updates. This results in incomplete learning dynamics, as the loss function struggles to balance between past and new data. Consequently, the model achieves a C-Index of 0.82, MAE of 155 days, KR of 0.63, and Fg of 0.16. While the replay mechanism demonstrates the ability to mitigate forgetting to some extent by reintroducing prior data, it lacks the parameter-level protection that EWC provides, leading to suboptimal retention of old information. This highlights the importance of combining replay with other CL techniques for robust knowledge preservation and minimal catastrophic forgetting.

#### CL EWC method apply Instead of Replay

In this setting, EWC is applied as the sole CL mechanism, removing the replay buffer entirely. EWC works by introducing a parameter regularization term in the loss function, which penalizes deviations from previously important weights. This helps retain critical features of earlier tasks by anchoring the model parameters to previously learned distributions. Due to that, the model achieves a C-Index of 0.83, MAE of 190 days, KR of 0.71, and Fg of 0.45. While EWC effectively minimizes forgetting by preserving parameter stability, its performance is limited in scenarios with significant distribution shifts in incoming data. Without replay, the model lacks the ability to refresh its understanding of earlier data, which can lead to over-reliance on parameter constraints and a higher MAE (see Table 1). This highlights the complementary nature of replay and EWC: while EWC stabilizes parameters for knowledge retention, replay reinforces memory by revisiting earlier examples, ensuring a more comprehensive learning process. The results emphasize the importance of combining these techniques to balance robust parameter protection and dynamic memory reinforcement, particularly in complex multimodal learning tasks.

This ablation study highlights the critical role of each component within the CL framework, underlining the significance of combining various mechanisms to achieve optimal performance. Our full framework, which integrates EWC, replay, cross-attention fusion, and XLNet-learned DNA embeddings, and CoxPH consistently outperformed all other model variants across key metrics, visualized in Fig 8.

**Fig 8.**
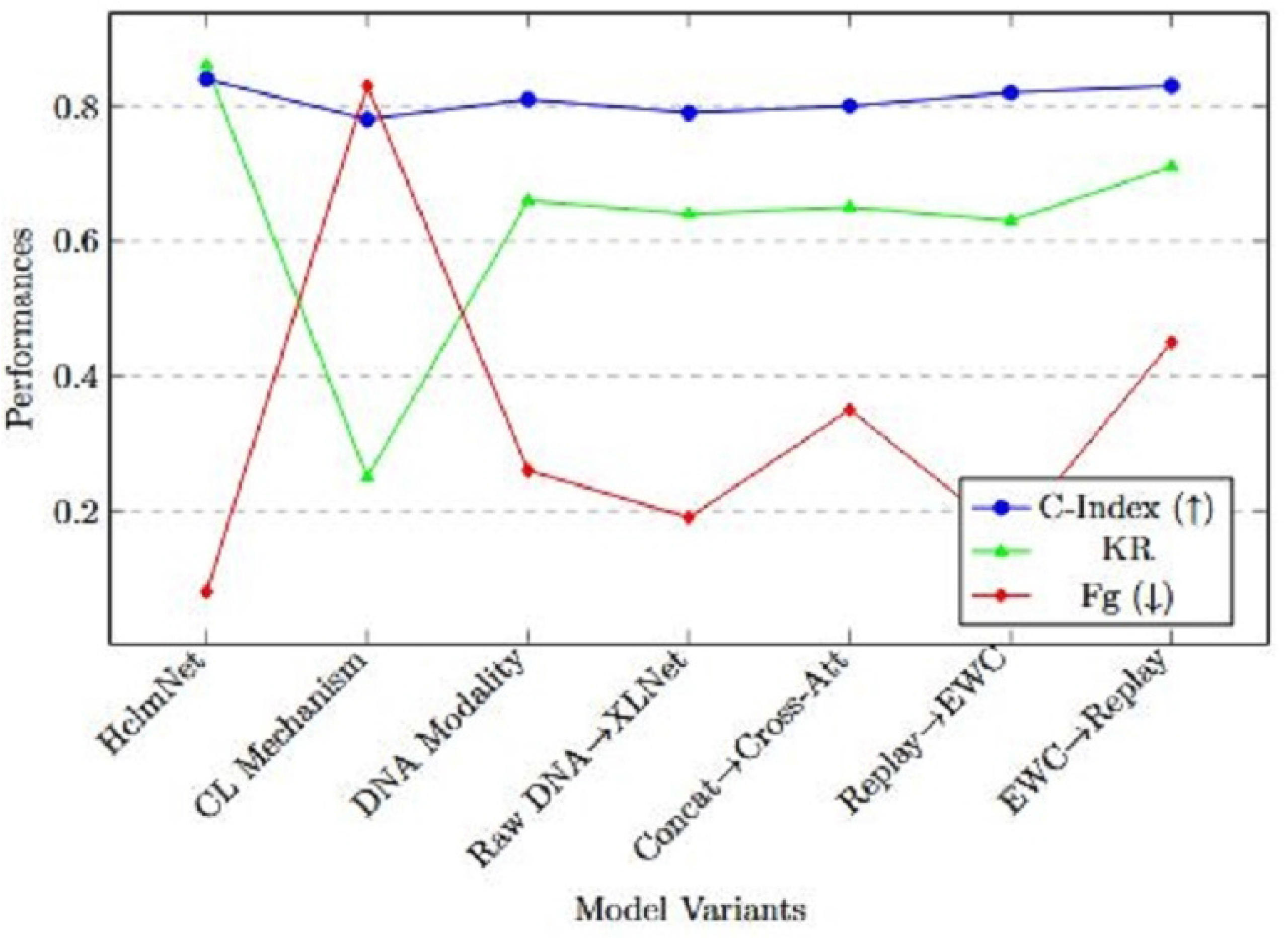
Ablation Study Results: Comparison of C-Index, KR, and Fg Across Variants. The proposed HCLmNet improves the C-Index by 7.7%, demonstrating better learnability. KR rises from 0.25 to 0.86, and Fg drops significantly by 90%, highlighting the effectiveness of combined replay and EWC strategies. Removing DNA modality and cross-attention notably affects performance, underscoring their importance.

Specifically, the inclusion of EWC and replay ensured a strong balance between knowledge retention and minimizing forgetting, while the cross-attention mechanism facilitated effective interaction between modalities. The integration of DNA embeddings contributed to the model’s ability to capture complex patterns from genetic data, strengthening its overall performance. In contrast, removing any of these components led to a noticeable degradation in model performance. For example, excluding the replay mechanism or using raw DNA features instead of embeddings resulted in higher forgetting and decreased knowledge retention, as observed in the increased MAE and reduced C-Index scores. These findings reinforce the importance of each individual design choice and demonstrate how the synergy of all techniques enhances the framework’s capability for incremental learning. Ultimately, the ablation study validates our approach, showcasing its robustness in retaining previously learned knowledge while adapting to new data without significant falling down in the performance.

### Main Results and Analysis

We conducted a comprehensive evaluation of our lung cancer survival prediction framework, leveraging three datasets: **D_3358P** for training and **D_200P** and **D_212P** for evaluation and CL. The base model was initially trained on the **D_3358P** dataset for 200 epochs using a batch size of 32, an Adam optimizer, a learning rate of 1 *×* 10^−4^, and a weight decay of 1 *×* 10^−4^. It achieved a training loss of **0**.**894** and a validation loss of **1**.**259**, as visualized in Fig 9. This indicates the model’s ability to identify patterns effectively during training, demonstrating convergence and stability.

**Fig 9.**
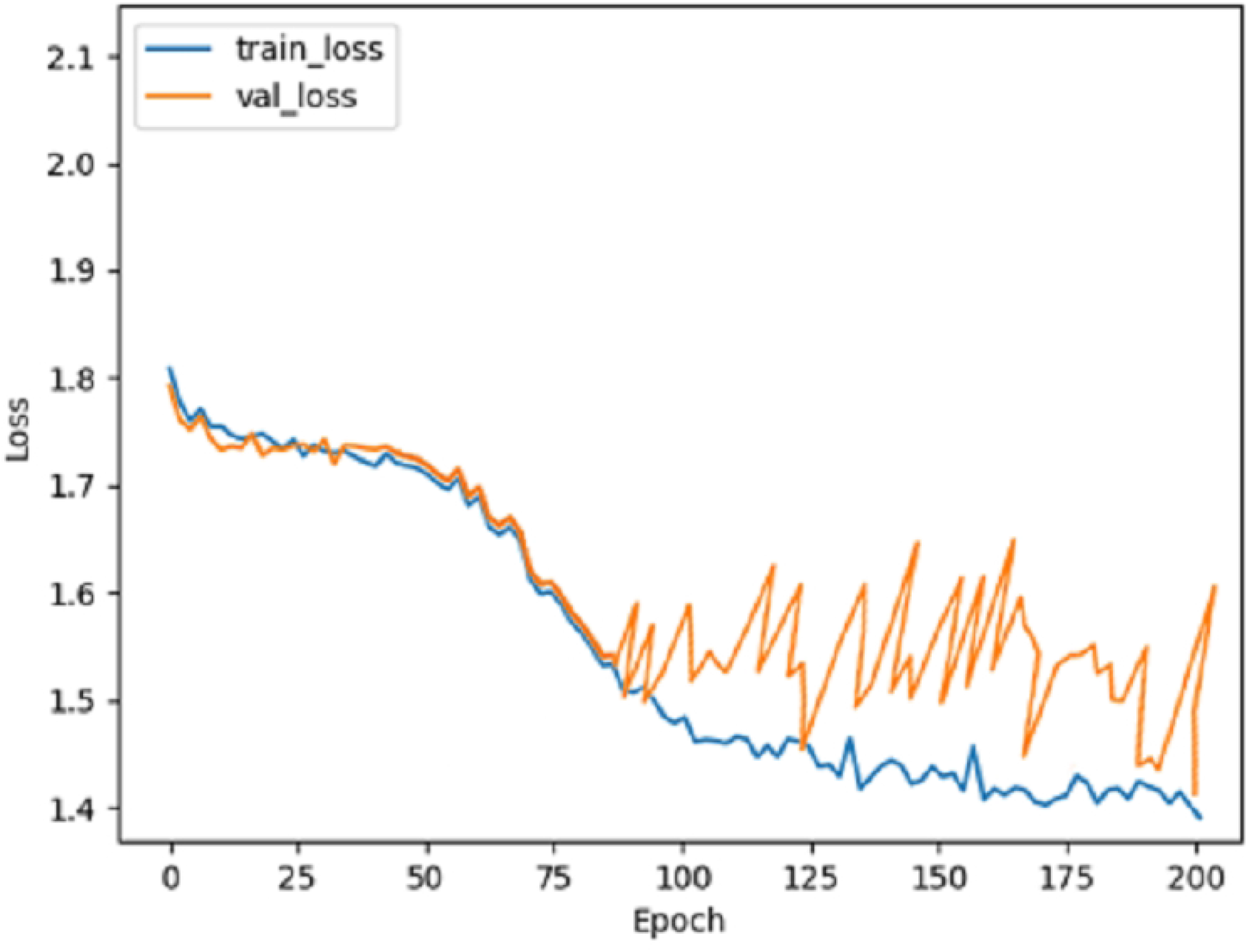
Baseline Model: Training and Validation loss over epochs.

Subsequently, the base model was evaluated on the **D_200P** dataset, comprising diverse lung cancer patients with varying ages and cancer subtypes. The evaluation yielded a **C-Index of 0**.**7656** and an **MAE of 189**.**4293**, which are reasonable given the complexity of predicting 5-year survivability. The survival probability graph in Fig 10, reflects this capability, showing a natural decline in survivability as the timeline progresses. This trend aligns with clinical observations, highlighting the model’s predictive reliability for a general lung cancer cohort. The evaluation thus validated the base model’s capability to generalize well across unseen data.

**Fig 10.**
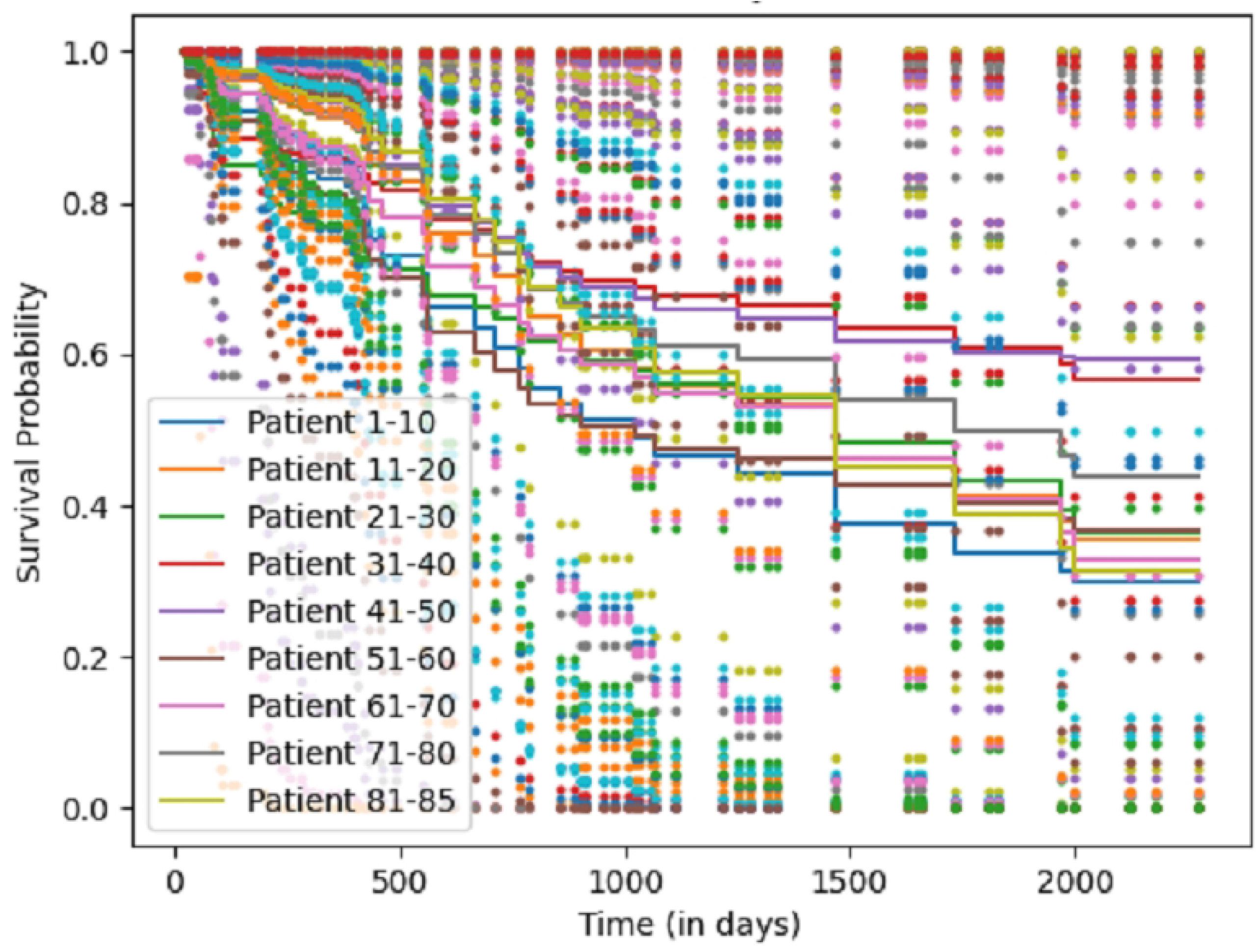
Baseline Evaluation: Survival probability trends over 5 years for 86 Patients. The plot shows the probability for 86 lung cancer patients over 5 years. Each point represents an individual patient’s survival prediction at specific time intervals, with distinct colors for each patient. The plot illustrates a realistic decline in survival probability over days.

To analyze the effectiveness of the CL framework, we introduced two incremental datasets: **D_200P** and **D_212P**. These datasets were incorporated sequentially into the pipeline to simulate real-world scenarios where new patient data continuously becomes available. The incremental learning module was designed to preprocess incoming data and leverage CL strategies to assimilate this knowledge while preserving previously learned patterns. The model underwent an additional 200 epochs of training on the combined dataset using the same configuration as the base training phase. This resulted in significantly improved metrics, with the training loss reduced to **0**.**0339** and the validation loss to **0**.**0324** (Fig 11). These low loss values suggest the model effectively adapted to new information without overfitting or degradation in performance.

**Fig 11.**
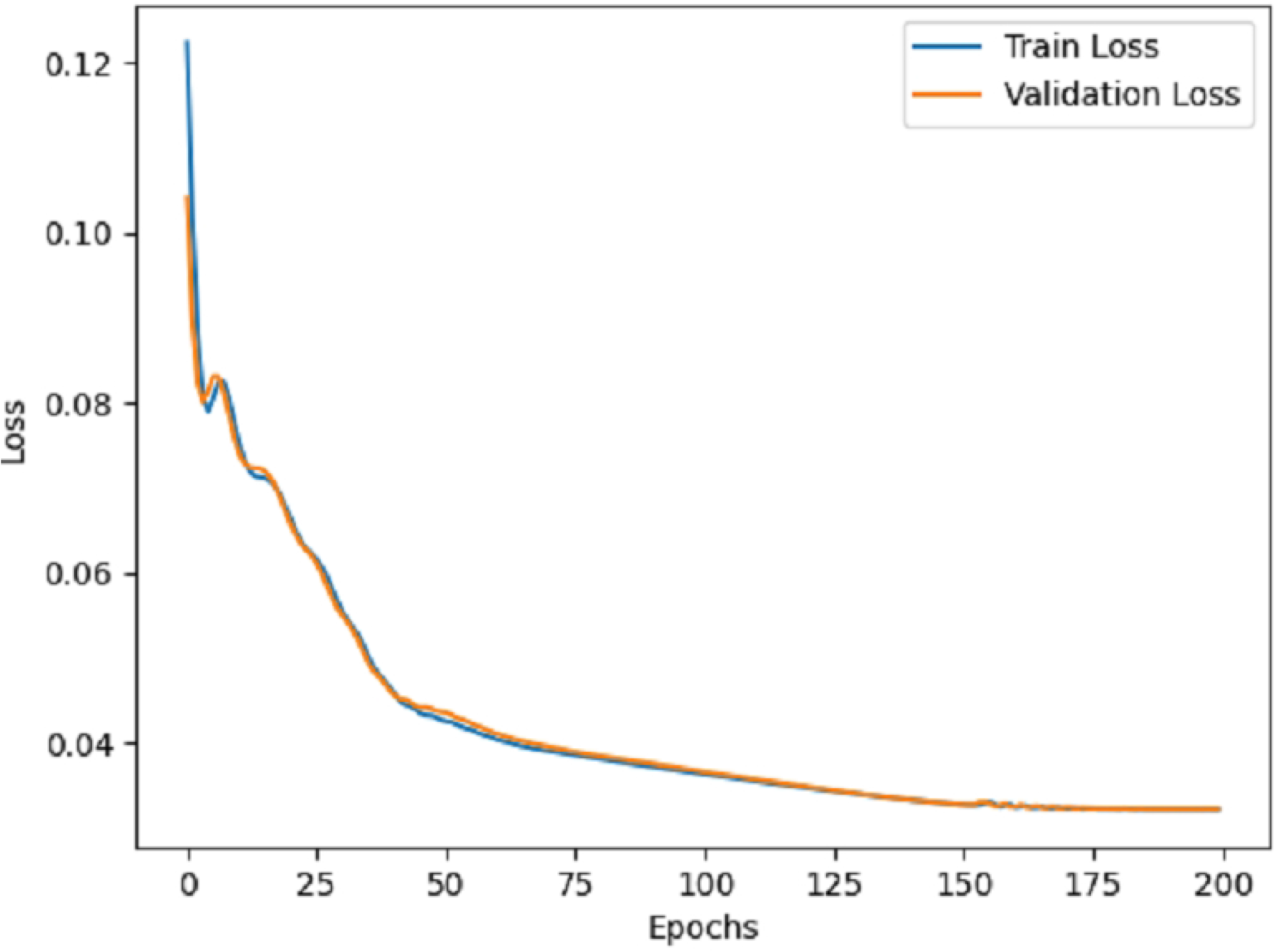
CL Model: Training and Validation loss over epochs.

In terms of predictive metrics, our HecmNet framework achieved a **C-Index of 0**.**8456** and an **MAE of 140**.**4233**, showcasing improved accuracy and discriminative ability compared to the base model. The survival prediction graphs, we plotted for patients over a 5-year timeline, exhibit smoother and more realistic trajectories with reduced variance between predicted probabilities and expected outcomes. This improvement is evident from Fig 12, where the survival probabilities for different patient groups more accurately align with clinical expectations. The CL framework effectively mitigated forgetting, preserved knowledge retention, and adapted to new data, ensuring that the model maintained high accuracy for both previously seen and newly introduced patients.

**Fig 12.**
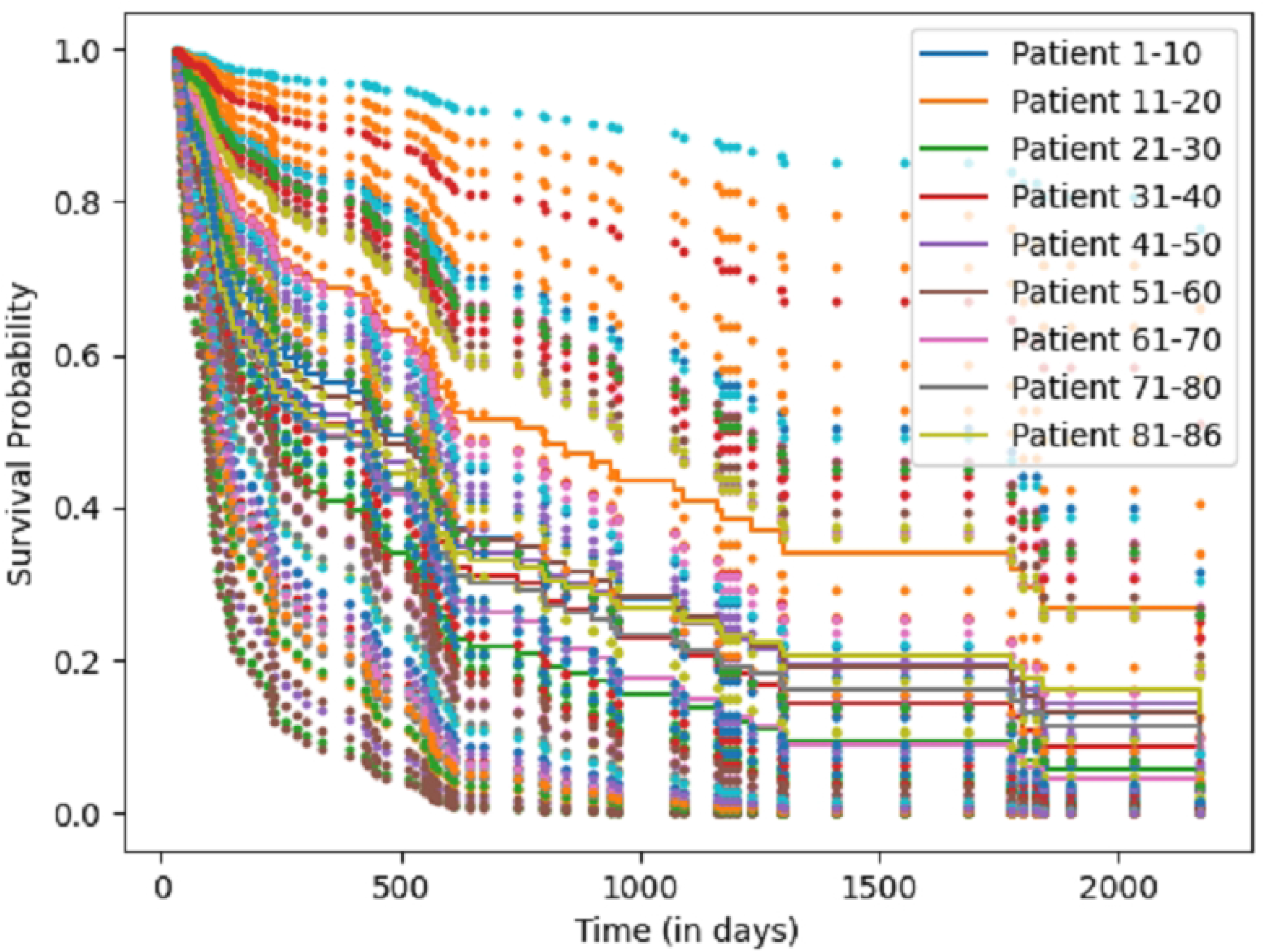
CL Evaluation: Survival probability trends Over time for 86 patients. The plot presents survival probability direction for 86 lung cancer patients, tracked over a period of 5 years. Each data point represents the survival prediction for an individual patient at specific intervals, with distinct colors indicating each patient’s unique trend. As new data is introduced and the model is trained with both previous and new data, the resulting evaluation shows smoother survival trajectories with reduced variability. This illustrates the effectiveness of the CL strategies in accurately predicting survival outcomes while minimizing noise and ensuring consistency across patient-specific survival paths.

Overall, these results emphasize the importance of each component within the framework. The base model demonstrated strong foundational capabilities, while the continual learning approach significantly enhanced performance by balancing knowledge retention and adaptation. This evaluation highlights the framework’s robustness, scalability, and practical applicability in predicting lung cancer survival outcomes, ensuring clinically meaningful predictions for a wide range of patients.

### Comparison with State of the Art Architecture

To assess our suggested model (HCLmNet) efficiency for lung cancer survival prediction, we compared it against several state-of-the-art survival models using the D 3358P and D 200P datasets. The results demonstrate that our model outperforms traditional and advanced methods in terms of both discriminative ability (C-index) and accuracy (MAE). Our HCLmNet achieved a C-index of 0.76 and an MAE of 189 days, significantly surpassing baseline models like CoxPH (C-index 0.65, MAE 247) and DeepSurv (C-index 0.67, MAE 252). Advanced models such as Trans-Surv, which integrates transformer-based survival modeling, reported a C-index of 0.71 and MAE of 258, falling short of our model’s results. Similarly, models incorporating cross-attention mechanisms, like Cross-Attention-LSTM (C-index 0.64, MAE 277), underperformed compared to HCLmNet, depicted in Table 2.

**Table 2.** Comparison of Recent State-of-the-Art Models for Lung Cancer Survival Prediction with base Model.

| Datasets Used: | D_3358P and D_200P |  |  |
| --- | --- | --- | --- |
| Model | Modality | C-index (↑) | MAE (↓) |
| CNNs-CoxPH | Clinical + CT + PET | 0.65 | 247 |
| DeepSurv | Clinical + DNA + CT + PET | 0.67 | 252 |
| RSF | Clinical + DNA + CT + PET | 0.52 | 250 |
| DeepHit | Clinical + DNA + CT + PET | 0.55 | 248 |
| FCN-SwinT-CoxPH | Clinical + DNA + CT + PET | 0.45 | 278 |
| Cross-Attention-LSTM | Clinical + CT + PET | 0.64 | 277 |
| Cox-Time | Clinical + DNA + CT + PET | 0.68 | 255 |
| Trans-Surv | Clinical + DNA + CT + PET | 0.71 | 258 |
| HCLmNet(Proposed) | Clinical + DNA + CT + PET | **0.76** | **189** |

**Notes:** C-index values closer to 1 indicate better discriminative ability, while lower MAE (**Day**) values indicate better accuracy in survival time prediction. The table compares models using different modalities to enhance lung cancer survival predictions over the traditional CoxPH baseline.

The superior performance of our model is attributed to its hybrid CL approach, which effectively leverages multiple modalities (clinical, DNA, CT, and PET) and balances retention of past knowledge with integration of new information. Additionally, the incorporation of cross-attention in the fusion layer optimizes feature alignment between modalities, ensuring robust survival predictions. This mechanism is crucial in enhancing the model’s understanding of patient-specific interactions, leading to improved discriminative power. Furthermore, the use of advanced loss functions, such as the CoxPH-based loss, aligns survival prediction with real-world clinical outcomes, contributing to the reduction in prediction errors (MAE). The results underscore HCLmNet’s robustness and adaptability, particularly when predicting survival probabilities over extended periods like five years.

In summary, our proposed HCLmNet establishes a new benchmark for survival prediction in lung cancer, delivering improved prediction accuracy and discriminative ability while addressing the limitations of traditional models.

### Assessment of Incremental Learning Strategies in Lung Cancer Survival Prediction

To evaluate the effectiveness of our CL strategies, we compared various state-of-the-art survival prediction models, focusing on their BP, NDP, NR, and Fg rates. The results demonstrate that our proposed HCLmNet outperforms other models across all CL metrics. HCLmNet achieves a high C-index of 0.84 and a remarkably low MAE of 140 days. Its BP (0.76) and NDP (0.83) highlight its ability to adapt to new data efficiently, while its superior KR (0.86) and minimal Fg (0.08) underscore the robustness of its hybrid CL mechanisms. The hybrid CL strategy we employed in HCLmNet combines EWC and replay-based approaches to balance plasticity and stability. EWC mitigates catastrophic forgetting by constraining updates to important weights, while the replay mechanism ensures retention of previously learned information by revisiting a curated memory buffer. Additionally, instance-level and class-level correlation modules enhance HCLmNet’s ability to align features across modalities (clinical, DNA, CT, and PET), improving the model’s overall stability and adaptability.

Fig 13 and Table 3 illustrate the CL evaluation, showing how HCLmNet maintains superior performance in baseline, new data, knowledge retention, and forgetting compared to other models. These findings highlight the effectiveness of the proposed framework in retaining critical knowledge and accurately predicting lung cancer survival.

**Table 3.** Evaluation of Continual Learning Approaches for Lung Cancer Prognostic Models.

| Datasets Used: | D.3358P, D.200P, and D.212P |  |  |  |  |  |
| --- | --- | --- | --- | --- | --- | --- |
| Model | C-index (↑) | MAE (↓) | Baseline Performance | NDP | Knowledge Retention | Forgetting (↓) |
| CNNs-CoxPH | 0.65 | 258 | 0.65 | 0.60 | 0.63 | 0.25 |
| DeepSurv | 0.70 | 252 | 0.70 | 0.68 | 0.67 | 0.19 |
| RSF | 0.72 | 250 | 272 | 0.70 | 0.69 | 0.45 |
| DeepHit | 0.75 | 248 | 275 | 0.73 | 0.72 | 0.23 |
| FCN-SwinT-CoxPH | 0.78 | 245 | 0.78 | 0.77 | 0.76 | 0.13 |
| Cross-Attention-LSTM | 0.77 | 244 | 0.77 | 0.76 | 0.75 | 0.17 |
| Trans-Surv | 0.76 | 204 | 0.76 | 0.74 | 0.73 | 0.15 |
| HCLmNet(Proposed) | **0.84** | **140** | **0.76** | **0.83** | **0.86** | **0.08** |

**Notes:** C-index values closer to 1 indicate better discriminative ability, while lower MAE (**Day**) values indicate better accuracy in survival time prediction. Forgetting (↓) represents the drop in performance on old data after learning new data (lower values are better).

**Fig 13.**
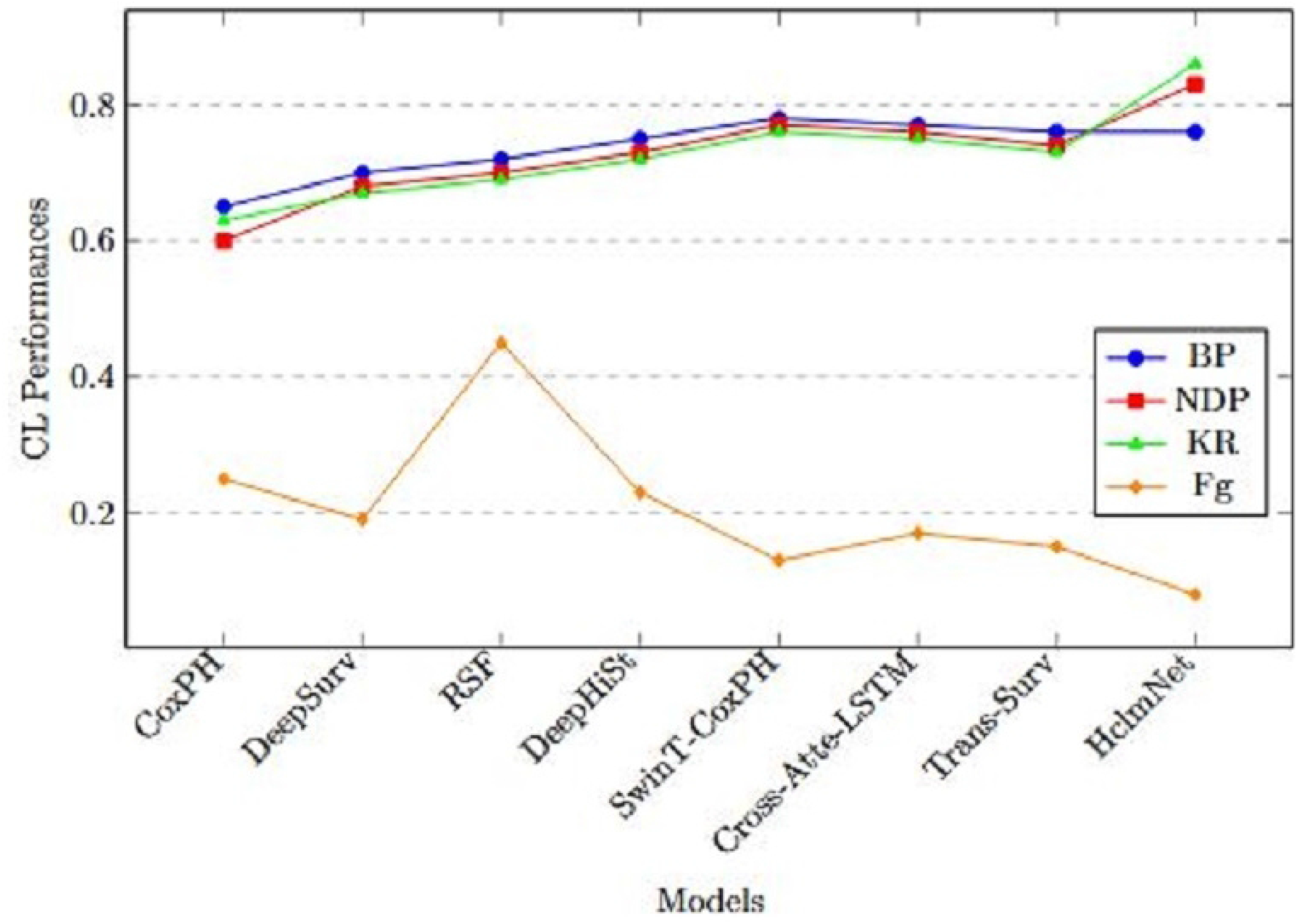
Continual Learning Evaluation: Baseline, New Data, KR, and Fg for Various Models.

### Computational Complexity Evaluation of Prognostic Models

In our experiments, we evaluated the computational complexity (CC) of various prognostic models for lung cancer using inference time (ms/sample) and FLOPS (G) as metrics, as summarized in Table 4. The CNNs-CoxPH model, with an inference time of 0.01 and 0.5 GFLOPS, was the most lightweight baseline but lacked the capacity to model complex multimodal interactions. DeepSurv and RSF required slightly higher computational resources at 0.1 and 0.05, with 3 and 2 GFLOPS, respectively, offering better multimodal data representation. DeepHit demonstrated increased complexity with 0.2 and 6 GFLOPS, reflecting the cost of incorporating advanced learning mechanisms. Models designed for domain-specific data, such as TransMIL (histology and genomics) and MOTCat (genomics and pathology), showed significantly higher computational demands, with TransMIL at 3.5 and 120 GFLOPS, and MOTCat at 4.0 and 140 GFLOPS, highlighting the cost of domain-specific feature extraction. The FCN-SwinT-CoxPH model, which integrates SwinT for multimodal feature extraction, required 5.0 and 250 GFLOPS, reflecting the computational expense of advanced feature integration. Similarly, the Cross-Attention-LSTM model achieved a balance between efficiency and complexity, with 2.0 and 100 GFLOPS, leveraging efficient data fusion mechanisms. Our proposed model, HCLmNet, exhibited the highest computational cost at 6.0 and 300 GFLOPS. This reflects its hybrid CL framework, combining replay-based memory and EWC with cross-attention mechanisms to ensure robust adaptation and accurate survival predictions. Despite its higher computational demands, HCLmNet offers exceptional potential for scenarios requiring robust multimodal integration and adaptive learning.

**Table 4.** Computational Complexity Evaluation of Prognostic Models for Lung Cancer.

| Datasets Used: | D.3358P, D.200P, and D.212P |  |  |
| --- | --- | --- | --- |
| Model | Modality | Inference Time (ms/sample) | FLOPS (G) |
| CNNs-CoxPH | Clinical + CT + PET | 0.01 | 0.5 |
| DeepSurv | Clinical + CT + PET | 0.1 | 3 |
| RSF | Clinical + CT + PET | 0.05 | 2 |
| DeepHit | Clinical + CT + PET | 0.2 | 6 |
| TransMIL [55] | Histology + Genomics | 3.5 | 120 |
| MOTCat [56] | Genomics + Pathology | 4.0 | 140 |
| FCN-SwinT-CoxPH | Clinical + DNA + CT + PET | 5.0 | 250 |
| Cross-Attention-LSTM | Clinical + CT + PET | 2.0 | 100 |
| HCLmNet (Proposed) | Clinical + DNA + CT + PET | 6.0 | 300 |

**Notes:** Inference Time refers to the time taken to process a single sample. FLOPS (G) represents the computational cost in billions of floating-point operations per second. Proposed model values are based on enhanced multimodal integration and CL efficiency.

## 7 Conclusion

This study introduces a hybrid CL multimodal framework designed to address the critical challenge of catastrophic forgetting in lung cancer survival prediction. By integrating EWC with replay-based strategies (ER, EICR, and ECCR), the framework adapts to new data while retaining prior knowledge. Leveraging SwinT-based feature extraction enhances the detection of critical features, such as ground-glass opacities and multiple tiny tumor instances with complex places, and XLNet-permutation effectively processes limited DNA datasets by learning latent data patterns. The proposed model outperforms state-of-the-art approaches, achieving a 7.7% improvement in C-Index (0.84), reducing MAE to 140, and significantly minimizing forgetting to 0.08. These results demonstrate its superior ability to deliver accurate and adaptive survival predictions in dynamic medical contexts. To further advance in this work, future research will focus on enhancing training efficiency through optimized DICOM image preprocessing and integrating federated learning to enable decentralized and scalable deployment in real-world healthcare environments. Finally, these findings signify a critical progression toward designing adaptive and dependable systems for predicting lung cancer survival outcomes.

## Data Availability

Dataset are publicly accessed in online. Link: https://aihub.or.kr/aihubdata/data/view.do?currMenu=115&topMenu=100&dataSetSn=228

## Acknowledgment

This work was supported by Innovative Human Resource Development for Local Intellectualization program through the Institute of Information & Communications Technology Planning & Evaluation(IITP) grant funded by the Korea government(MSIT)(IITP-2024-RS-2022-00156287, 40%). This work was supported by Institute of Information & communications Technology Planning & Evaluation (IITP) under the Artificial Intelligence Convergence Innovation Human Resources Development (IITP-2023-RS-2023-00256629, 30%) grant funded by the Korea government(MSIT). This research was supported by the MSIT(Ministry of Science and ICT), Korea, under the ITRC(Information Technology Research Center) support program(IITP-2024-RS-2024-00437718, 30%) supervised by the IITP(Institute for Information & Communications Technology Planning & Evaluation)

